# Analysis of multiple causes of death: a review of methods and practices

**DOI:** 10.1101/2022.08.01.22278086

**Authors:** Karen Bishop, Saliu Balogun, James Eynstone-Hinkins, Lauren Moran, Melonie Martin, Emily Banks, Chalapati Rao, Grace Joshy

## Abstract

**Background:** Research and reporting of mortality indicators typically focus on a single underlying cause of death selected from multiple causes recorded on a death certificate. The need to incorporate the multiple causes in mortality statistics - reflecting increasing multimorbidity and complex causation patterns - is recognised internationally. This review aims to identify and appraise relevant multiple cause analytical methods and practices.

**Methods:** We searched Medline, PubMed, Scopus and Web of Science from inception to December 2020 without language restrictions, supplemented by consultation with international experts. Eligible articles included those analysing multiple causes of death from death certificates. The process identified 4,080 articles; after screening, 434 full texts were reviewed.

**Results:** Most reviewed articles (77%, n=332) were published since 2001. The majority examined mortality by “any-mention” of a cause of death (87%, n=377) and assessed pairwise combinations of causes (56%, n=245). Recently emerging (since 2001) were applications of methods to group deaths based on common cause patterns using, for example, cluster analysis (2%, n=9), and the application of multiple cause weights to re-evaluate mortality burden (1%, n=5). Multiple cause methods applied to specific research objectives are described for recently emerging approaches.

**Conclusion:** This review confirms rapidly increasing international interest in the analysis of multiple causes of death and provides the most comprehensive overview of methods and practices to date. Available multiple cause methods are diverse but suit a range of research objectives, that with greater data availability and technology could be further developed and applied across a range of settings.

## Introduction

Mortality statistics are crucial to population health as they provide fundamental information about health status, disease aetiology, trends, and patterns of diseases in different populations. They inform health services, health policy development and planning, as well as research and can be used to evaluate the impact of health intervention programs.^1,2^ Therefore, it is critical that accurate and reliable information about the diseases and health conditions that cause death are appropriately analysed.

Death typically results from the interplay between multiple health conditions. The standard international format of the death certificate (Figure 1) facilitates recording the certifying doctor’s medical opinion of all diseases and conditions involved in the death including the underlying and non-underlying (intermediate and immediate) causes in Part I, and significant other contributing causes in Part II. If the certificate is completed correctly, the underlying cause (UC) reflects the initiating condition, that is, one that could be avoided by some preventative mechanism to interrupt the sequence leading to death. The medical certification process reflects the multifaceted pathological processes leading to death. However, the recording of a single disease as the UC can be complex and misclassification of the UC can occur when several causal pathways are involved.^3-8^ Following the medical certification of the cause of death, an international coding standard is applied to all causes reported on the death certificate to endorse the reported UC or select a more appropriate alternative to be used for statistical reporting (including international comparisons) and epidemiological studies.^9^ Deaths data thus contain the standardised UC and all other causes that were involved in the death (associated causes).

**Figure 1.**
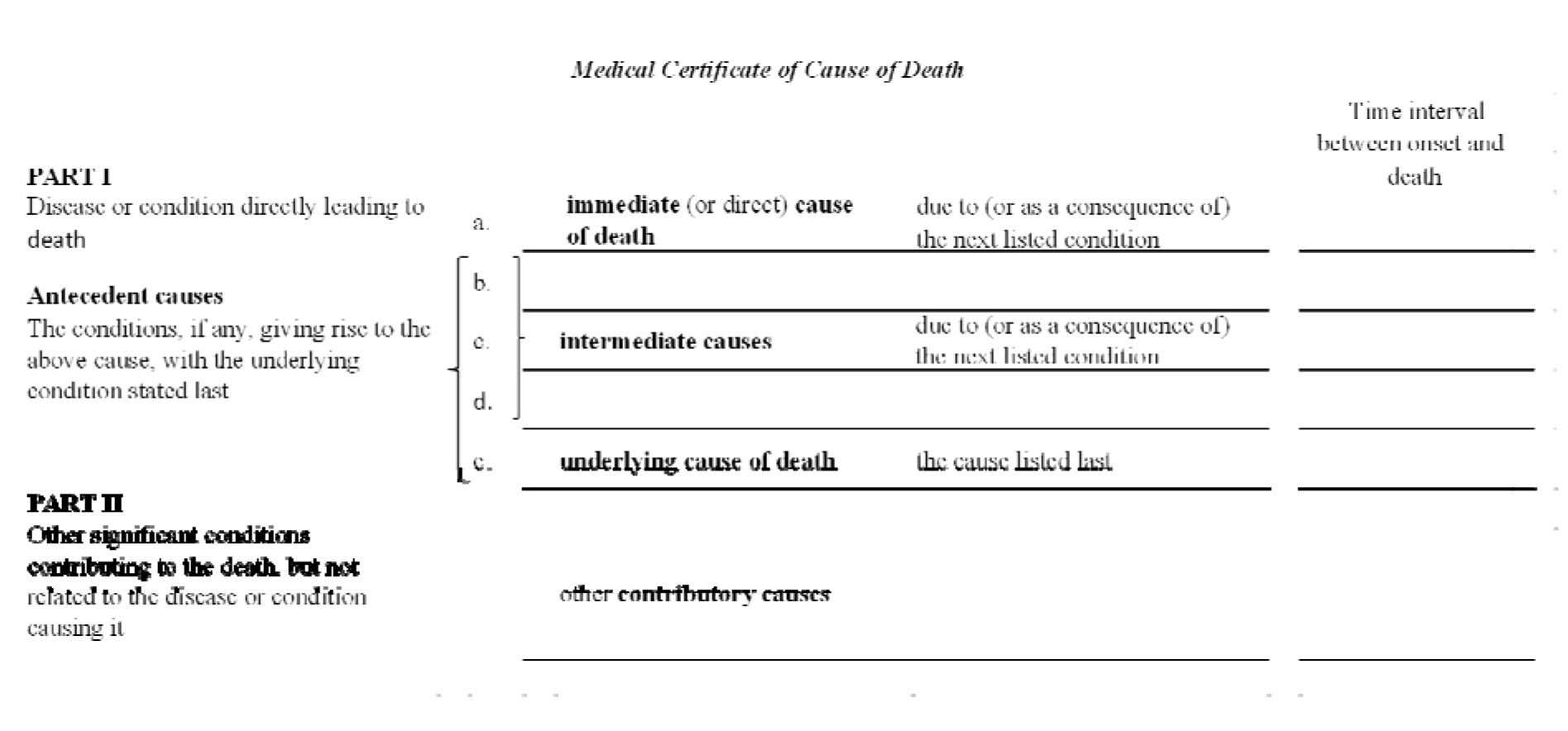
Layout of a standard international form of Medical Certificate of Cause of Death.

Despite the vast amount of information that is collected about the causes of deaths, mortality statistics typically use only the (single) UC. Researchers have long recognised that the UC alone does not adequately describe the pathological processes responsible for most deaths,^10-12^ and potentially understates the importance of other significant contributing causes of death.^1,13^ International support for the need to assess the multiple causes of death (MC) to complement statistics based on the UC approach is well established but methods used are diverse. To adequately inform population health initiatives, it is essential that all diseases and conditions contributing to death receive appropriate attention.

Previous studies^10,14-16^ have attempted to appraise and summarise the assortment of methods used to measure the involvement of all causes on the death certificate, however they are limited in scope and were performed over a decade ago thus not capturing recent methodological advances in the analysis of MC. This review, supplemented by consultations with subject experts, aims to identify and appraise the methods used in analyses of MC data, providing a comprehensive and up-to-date account of methods and practices that are used to describe and measure the involvement of multiple health conditions in causing death.

## Methods

### Search strategy

To identify articles that analyse MC data, we searched the Medline, PubMed, Scopus and Web of Science databases, each from inception to 31 December 2020, for original research without any restriction on language or country of study. To maximise the number of relevant articles related to multiple causes of death, we included search terms covering ‘multiple*’ AND ‘cause*’ AND (‘death’ OR ‘mortality’) and other variations such as (‘associated’ OR ‘contributory’ OR ‘underlying’) AND ‘cause*’ (Supplementary File 1).

### Selection of articles

Two authors (KB and SB) each independently screened 50% of titles and abstracts of the identified articles, beginning with a random sample of 5% articles in duplicate. Disagreements were solved by consensus.

### Eligibility criteria

Articles were eligible if they: reported using death registration or death certificate data and applied a multiple cause method to calculate a multiple cause indicator; or derived a measure of mortality based on the multiple causes of death. We considered all study designs, except case reports, case series and forensic reports. Research that used only the UC for analysis, or that used MC data but did not apply or report a measure based on multiple causes were excluded. Articles based on verbal autopsy, narrative, reviews, and non-peer reviewed literature were also excluded.

### Data extraction

For the included articles, study characteristics (authors’ names, journal name, year published, study design, study period, country, source of multiple cause data and main cause of interest) and decedent characteristics (age, sex and number of deaths evaluated) were extracted using a full text review. Each article was categorised into one or more categories according to the objectives of the application of MC methods as articles that: described cause-related mortality based on ‘any-mention’ of a cause; assessed the joint involvement of causes according to pairwise disease occurrence on death certificates; described mortality for clusters of >2 commonly co-occurring causes; and measured cause-related mortality burden by weighting multiple causes.

### Audit of experts

To identify unpublished methods in practice, the search strategy was supplemented by consultation with subject experts. Contacts were identified from affiliations of relevant papers and recommendations from experts in the field, and included representatives from agencies such as the Multiple Causes-of-Death Network (https://mcod.web.ined.fr), the World Health Organization Family of International Classification collaborating centres, and national statistical offices of countries, including the United States, England, Canada, New Zealand, Italy and Australia. During December 2020, emails were sent to 261 contacts asking regarding their awareness of relevant studies, particularly recently accepted or unpublished papers or reports that used multiple cause methodology. We received 41 responses resulting in a response rate of 15.7%.

### Statistical methods

Articles included in full text review were classified based on *a priori* selected mutually exclusive categories of statistical methods used to analyse multiple causes of death as: methods based on any-mention; methods to assess pairwise occurrence of causes; methods based on groups of >2 co-contributing causes; and methods based on weighting of multiple causes. Following full text review, included articles published from 2015 onwards were classified based on the main research objective into four categories: describe cause-related mortality; identify co-contributing causes; assess relationships between co-contributing causes; assess impact of risk factors; and other objectives. For articles published between 2015 and 2020, we mapped the research objectives against the MC methods that were used to achieve them. We did not evaluate publication bias as this review focussed on methodological practices applied in each article rather than on results.

## Results

Overall 8,070 articles were identified: 8,002 from the database search and 68 from responses to the consultations. After removing duplicates, 4080 articles were selected for title and abstract screening (Figure 2). Due to the large number of articles and limited resources, 101 articles that appeared to only use methods based on any-mention were excluded without full text review. Four review articles were identified in the screening, and a manual search of these identified a further 25 potentially eligible articles. A total of 602 articles were selected for full text review, including 25 from the consultations (Figure 2). From these, 434 articles were included, and the multiple cause methods applied in each were assessed. The remaining 168 articles did not meet the inclusion criteria: four were reviews, 61 were not in scope, 53 were irretrievable (including 13 in a foreign language), 28 were communications or conference abstracts and 22 were non-peer reviewed articles (none of them employed methods other than those *a priori* identified) and were excluded from full text review.

**Figure 2.**
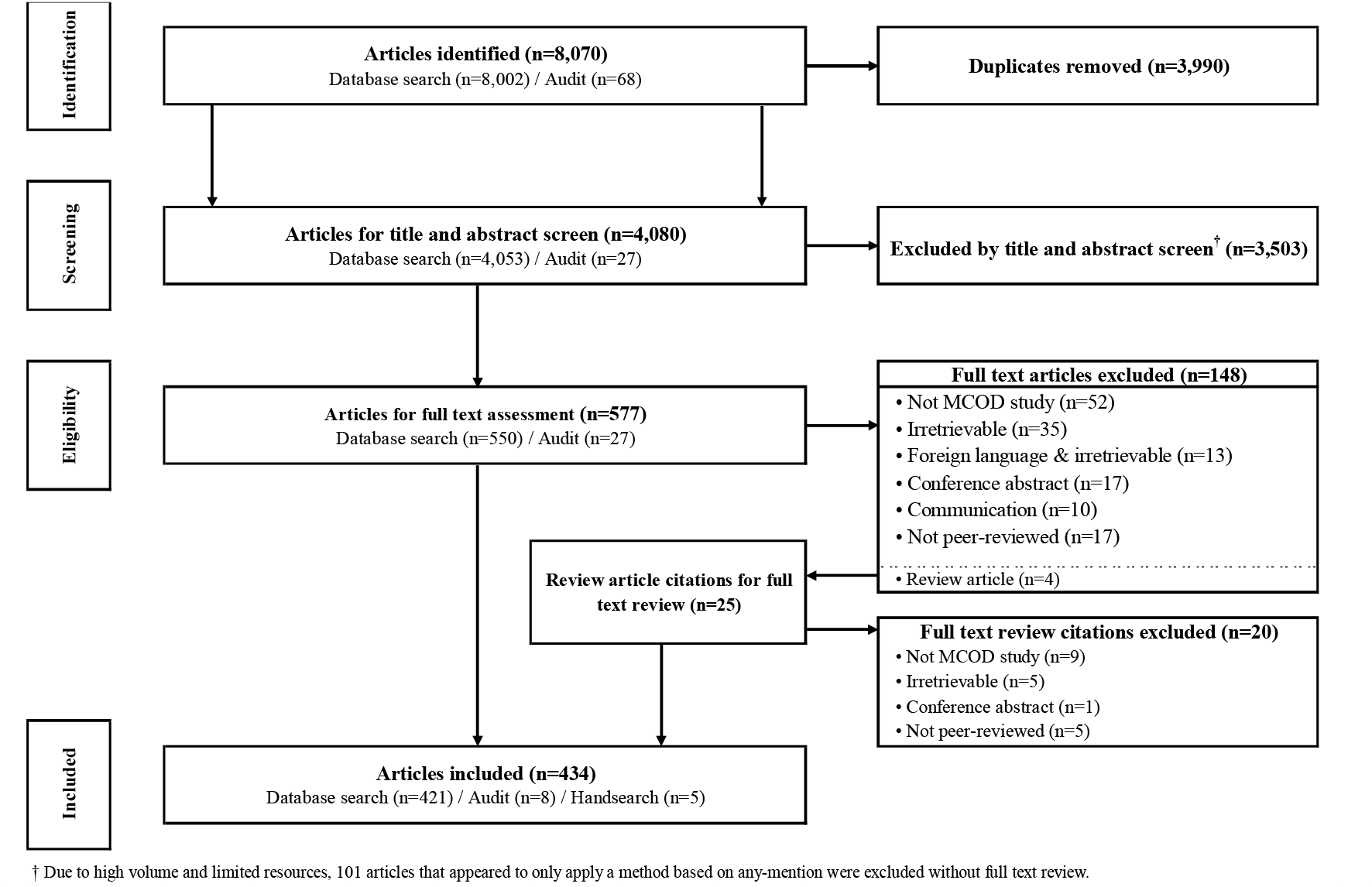
Study flow diagram.

A summary of the characteristics of included articles is presented in Table 1. The number of articles using MC methods increased over time; more than three-quarters (n=332, 76.5%) were published after 2001 (Table 1). Most articles assessed deaths registered in the United States (n=222, 12.9%), Brazil (n=47, 2.7%) and the United Kingdom (n=43, 2.5%) (Table 1). In most countries, the application of MC methods rose over time, with notable increases during 2001–2020 (Figure 3). Infectious diseases (largely HIV/AIDS) were the most common cause of interest in the application of MC methods (n=76, 17.5% articles), followed by external causes (n=69, 15.9% articles) of which most assessed drug-related deaths. Population-level analysis of all causes of death using multiple cause methods were found in 44 (10.1%) articles. Cross-sectional evaluation of deaths was the most common study design (n= 353, 81.3% articles). Most articles reviewed were in English (n=416, 95.9%) and Portuguese (n=13, 3.0%).

**Table 1.** Characteristics of articles included in the study.

|  | **Number** | **Percent** |
| --- | --- | --- |
| **Total number of articles** | **434** |  |
| **Year published** |  |  |
| 1980 or earlier | 19 | 4.4 |
| 1981-1990 | 29 | 6.7 |
| 1991-2000 | 54 | 12.4 |
| 2001-2010 | 114 | 26.3 |
| 2011 to present | 218 | 50.2 |
| **Country of data<sup>1</sup>** |  |  |
| United States | 222 | 12.9 |
| Brazil | 47 | 2.7 |
| United Kingdom | 43 | 2.5 |
| Italy | 30 | 1.7 |
| France | 23 | 1.3 |
| Sweden | 19 | 1.1 |
| Australia | 19 | 1.1 |
| Canada | 7 | 0.4 |
| Finland | 7 | 0.4 |
| Norway | 6 | 0.3 |
| All other countries | 50 | 2.9 |
| **Research Focus<sup>1</sup>** |  |  |
| Population level analysis of all causes | 44 | 10.1 |
| Specific causes of death |  |  |
| Infectious diseases | 76 | 17.5 |
| *HIV/AIDS* | 22 | 5.1 |
| *Hepatitis* | 9 | 2.1 |
| *Sepsis* | 8 | 1.8 |
| External causes | 69 | 15.9 |
| *Drug-related* | 23 | 5.3 |
| *Alcohol-related* | 15 | 3.5 |
| Cardiovascular diseases | 50 | 11.5 |
| Endocrine diseases | 43 | 9.9 |
| *Diabetes* | 37 | 8.5 |
| Neurological diseases | 35 | 8.1 |
| Cancer | 25 | 5.8 |
| Respiratory diseases | 23 | 5.3 |
| Digestive diseases | 13 | 3.0 |
| Musculoskeletal conditions | 12 | 2.8 |
| Mental conditions | 11 | 2.5 |
| All other causes | 33 | 7.6 |
| **Study design** |  |  |
| Cross-sectional | 352 | 81.1 |
| Cohort / longitudinal | 69 | 15.9 |
| Case control | 7 | 1.6 |
| Other | 6 | 1.4 |
| **Language of article** |  |  |
| English | 416 | 95.9 |
| Portuguese | 13 | 3.0 |
| Other | 5 | 1.2 |

<sup>1</sup>. Numbers may not sum to the total as multiple categories apply.

**Figure 3.**
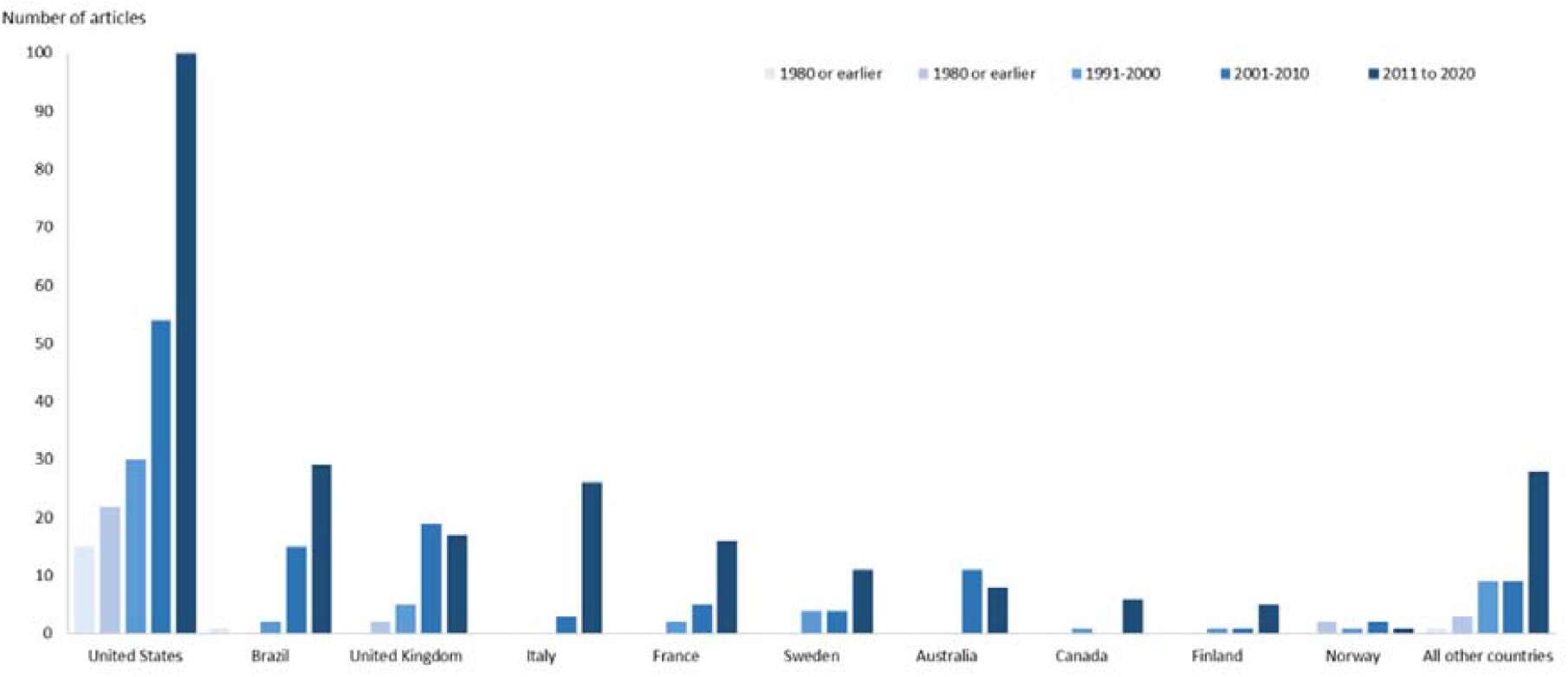
Number of articles by country and year of publication.

Articles were broadly categorised by the methods applied in analysing MC as those describing mortality based on any-mention of a cause; examination of pairwise occurrence of diseases on death certificates; assessment of mortality based on groupings of >2 co-contributing causes; and calculation of cause-related mortality burden based on weighted multiple causes. The results for each category are described in detail below.

### Descriptive measures of multiple causes based on any-mention

The full text review identified 86.9 % (n=377) articles that assessed cause-related mortality based on any-mention of the cause (Table 2, Supplementary File 2). The methods applied in these articles included basic summary (or univariate)^17^ statistics applied to a specific index cause. Examples of specific methods include multiple cause indicators that describe the number of causes (*n*) involved in each death, the average number of causes per death and frequency or percentage distributions of *n*, each illustrating the extent to which multiple causes occur in the deaths data. Also included here were articles that aimed to evaluate cause-related mortality using rates based on any-mention of a cause; that is, by counting each death that mentions the cause of interest anywhere on the death certificate.^18-21^ Some evaluated the leading causes of death using any-mention^22-27^ (to understand the most common causes involved in deaths. Assessment of temporal trends in any-mention rates against rates based on the UC were used to highlight changes in certification or coding practices and changing patterns of disease contribution to death.^28-30^ Comparisons between countries in multiple cause indicators can be used to emphasise differences in certification practices between countries.^3,31-33^

**Table 2.** Summary of approaches to assess multiple causes of death in included studies.

| Methodological approach | Example articles<sup>1</sup> |
| --- | --- |
| **1. Methods based on any-mention (n=377, 86.9%)**<br>**Uses:** to assess the extent of multiple causes and compare measures based on UC to measures based on MC<br>**Examples:** average number of causes; rates by ‘any-mention’; comparison of underlying to multiple causes (e.g. SRMU)<br>**Strengths:** identifies causes less visible using UC approach<br>**Limitations:** metrics based on any-mention inflate mortality estimates as deaths are counted more than once | Wall (2005)<br>Goldacre (2006)<br>Desesquelles (2014)<br>Goldberger (2015)<br>Zoppini (2018)<br>Simmons (2019)<br>Sampaio (2020)<br>Cano (2020) |
| **2. Methods to assess pairwise occurrence of causes (n=245, 56.5%)**<br>**Uses:** to identify and assess co-contributing causes of death<br>**Examples:** leading underlying causes for a specific associated cause; mortality odds ratios for the presence of specific causes<br>**Strengths:** identifies most commonly occurring comorbid conditions at death; measures the strength of association between two co-contributing causes on the death certificate (e.g. odds ratio, CDAI)<br>**Limitations:** does not measure causality; can overlook relationships between two non-underlying causes | Redelings (2005)<br>Rockett (2007)<br>Redelings (2007)<br>Desesquelles (2012)<br>Duncan (2014)<br>Chazal (2018)<br>Turner (2018)<br>Quast (2020) |
| **3. Methods based on groups of >2 co-contributing causes (n=9, 2.1%)**<br>**Uses:** to identify frequently co-occurring causes and assess mortality trends due to grouped causes<br>**Examples:** combine causes based on known risk associations or cluster analysis, social network analysis and data mining according to patterns in multiple causes<br>**Strengths:** identifies highly correlated diseases and risk conditions among large datasets<br>**Limitations:** data-driven methods are difficult to replicate or apply across different settings | Stallard (2002)<br>Frova (2009)<br>Yoon (2011)<br>Barbieri (2017)<br>Hassanzadeh (2017)<br>Jiang (2017)<br>Egidi (2018)<br>Villela (2018)<br>Adair (2020) |
| **4. Methods based on weighting of multiple causes (n=5, 1.1%)<sup>2</sup>**<br>**Uses:** to re-evaluate mortality metrics based on weighted multiple causes<br>**Examples:** ascribing fixed or variable weights to each cause such that the weights in each death sum to one<br>**Strengths:** can prescribe causal responsibility to selected diseases (e.g. antecedent conditions); incorporates a mortality contribution of all relevant conditions<br>**Limitations:** difficult to ascribe causal responsibility across cause of death data so weights are arbitrary | Piffaretti (2016)<br>Moreno-Betancur (2017)<br>González (2019)<br>Breger (2020)<br>Xie (2020) |

<sup>1</sup> See Supplementary Appendix 2 for full reference
<sup>2</sup> Articles that applied multiple cause weighting methods and data-driven methods for grouping deaths based on multiple cause patterns were all applied since 2016

Rate ratios were commonly applied to indicate the extent to which a cause occurs as the UC versus non-UC. A common approach compared the occurrence of the cause as any-mention to its occurrences as the UC (the approach varies according to whether counts or rates are used in the ratio and whether the any-mention versus non-underlying mentions are compared to the UC.^10^ One form of this measure, the standardised ratio of multiple to underlying causes (SRMU),^13,34^ encourages a harmonised approach to calculating this indicator; here the rate ratio is calculated as the age-standardised rate for any-mention of the cause compared to the age-standardised rate when the cause is the UC. This indicator describes the extent to which the cause is selected as non-UC relative to UC (with 1 indicating the cause is always the UC, 2 indicating equal representation as UC and non-UC and >2 indicating the cause is more often a non-UC). Country comparisons of this method can be used to assess variation in certification.^32^

Articles that used counts or rates of any-mention of causes to derive other summary measures of mortality, for example potential years of life lost^35^ and life expectancy^36-41^ were also categorised here. Further examples of articles that applied these methods are in Table 2.

### Assessing pairwise contribution to mortality

Articles that investigated the relationships between two causes of death reported on the same death certificate were categorised here; we found 56.5% (*n*=245) articles in this review (Table 2, Supplementary File 2). A distinguishing feature of these methods is that the joint frequencies of just two causes are the focus of the analysis. To investigate relationships between two causes, researchers assessed the involvement of a specific UC with one or more associated causes or vice versa (e.g. the nature and spread of an associated or immediate cause for an UC of interest). Typically, the objective is to understand which comorbid conditions commonly occur with a specific cause using frequency or percentage distributions of the most common associated causes for a specific UC and vice versa.^42-45^

More comprehensive applications of odds ratios (mortality odds ratios) and relative rates was found in articles that aimed to measure the associations between comorbid medical conditions involved in the death for a specific cause of interest. Mortality odds ratios were used to assess the odds of any-mention of a specific cause with other comorbidities at death. ^9,35,46-57^ Rate ratios were used to assess mortality burden according to whether another specific cause was present or absent, or to deaths in general.^58,59^ In most cases the application of mortality odds ratios disregards the role of the cause, thereby enabling relationships between two non-UCs to be included in the assessment. Previous reviews of measures of association for multiple cause of death discuss the applicability of several approaches including: matched mortality odds ratios, matched exposure odds ratios,^15^ and Yules Q, Positive Matching Index, Forbes’ coefficient and, Wise and Sorvillo ratio),^14^ concluding the most suitable to be those that do not consider non-matches.

A more recently introduced measure, the ‘cause of death association indicator’ (CDAI) compares the standardised rate of involvement of a cause of interest reported together with a specific UC to its involvement in death overall (that is with any UC).^13,34,60^ The CDAI aids understanding whether a non-UC of interest is more common with a specific UC than with all UCs combined. In this indicator, the role of the causes is fixed thereby requiring reverse comparisons for each UC and non-UC of interest.

### Assessing mortality patterns for grouped causes of deaths

Articles categorised here aimed to assess mortality from either a cluster of decedents where the grouping is based on patterns in the multiple causes or by known disease associations, or by a cluster of specific causes grouped according to some measure of ‘similarity’ or closeness. These approaches were applied in 2.1% (n=9) articles and were considered as newly emerging methods, with all but one article published since 2009 (Table 2, Supplementary File 2). Methods for grouping multiple causes of death were largely used to understand more complex relationships between multiple causes of death or to highlight patterns of disease that commonly co-contribute to death with more than two causes of interest. In some articles, the groupings were user-specified combinations of causes^11,61-63^ while others used data-driven methods such as cluster analysis,^64,65^ social network analysis^66^ and more exploratory methods of data mining.^67,68^ Social network analysis identified links (and their strengths) between causes of death, cluster analysis methods grouped decedents on the basis of similarity between causes, that is, based on the causes of death that commonly co-contribute to death, and data mining techniques were applied to identify complex patterns in mortality data^68^ and assess temporal evolution of the leading clusters of conditions that cause death.^67^

### Multiple-cause weighting methods

We identified five (1.1%) articles that aimed to calculate cause-related mortality by ascribing weights to each cause in the death record^1,69-72^ (Table 2, Supplementary File 2). The multiple cause weighting methods identified in this review, assigned weights to each cause such that within each death, the weights summed to 1.0. With this approach, the counting unit (deaths) is preserved enabling measures of cause-related mortality (rates, years of life lost, etc) to be recalculated based on the weighted counts of deaths. The included articles illustrated their proposed method by estimating socioeconomic inequalities in mortality,^69^ conditions whose contribution to death is underestimated,^1,70^ proportional mortality,^72^ and the relative risk of cause-specific mortality among individuals with human immunodeficiency virus versus those without.^71^

Various strategies were noted for ascribing weights, for example, weighting all causes equally as 1/*n* where *n* is the number of causes involved in the death, weighting the UC a fixed amount (e.g. 50%) with the remaining amount (in this case 50%) apportioned equally across the other causes, or weighting the UC twice that of other causes. The strategies also varied according to the causes included for weighting – all causes versus the UC plus contributing causes from Part II of the death certificate.

Among included texts, 45.7% (n=198) applied >1 MC method; most frequently (n=187) an any-mention method and a pairwise assessment of causes of death (Table 3). Four of five applications of weighting methods also reported indicators using any-mention.

Across all applications of MC methods, the techniques varied in regard to 1) whether the study included a comparison of multiple to underlying causes; 2) whether the study used all non-UCs, or a specified set based, for example, on the location on the death certificate (Part I or Part II); 3) whether ill-defined or external causes of death were considered; and 4) whether duplicate mentions of the cause of interest were excluded.

### Research Objectives

The objectives for applying multiple causes analyses varied. Some broad themes were identified in the aims: to describe cause-related mortality using MC; to identify co-contributing causes (that is, the associated causes for a specific UC and vice-versa); to assess relationships between causes using a measure of association; or assess the impact of risk factors on mortality. A residual category of ‘other’ objectives captured articles that intended to measure the contribution of all causes of death in a population using MC methods and those with aims that centred around ‘exploring’ the MC data. In this subset of articles published between 2015 and 2020 (n=133), most (54.9%, n=73) aimed to describe the cause of interest using multiple causes of death and applied methods based on any-mention in 66 articles and pairwise assessment in 39 (Table 4). Of the 28 articles whose main aim was to identify co-contributing causes for a specific cause of interest, nearly all applied methods based on any-mention (n=24) and pairwise assessment of causes on the death certificate (n=25) with only 1 that grouped more than two causes for analysis. Most notable, were the applications of emerging approaches to analysing multiple causes. Foremost were whole of population analyses to quantify the contribution of all causes that contributed to death by weighting each one as described above.^1,69^ Though weights were arbitrary, these novel methods lend to capturing all causal contribution at a population level, which by restricting the selection of causes can convey the mortality contributed by antecedent causes^1,69^ or if required, only the complications of diseases for targeted prevention efforts.^73^ Other examples of recently emerging methods assessed the impact of risk factors by quantifying the contributing causes of death (using weighting) among known AIDS cases for comparison of causes of death by exposure to injecting drug use,^71^ assessing the impact on mortality trends of deaths reporting the presence of conditions known to be associated with obesity,^61^ and by using cluster analysis techniques to assess mortality differentials between clusters of discrete groupings of causes associated with obesity.^64^

## Discussion

Our review and audit confirm rapidly increasing international interest in the use of multiple causes of death data in mortality research and provides the most comprehensive overview of methods and practices to date. Most articles identified were published since 2011 and were from countries with health information management systems enabling collection and recording of multiple causes of death; the United States, Brazil, the United Kingdom, Italy, and France being the most common. Articles analysing multiple causes of death were highly skewed towards those using descriptive measures applied to any-mention of a cause of death and those assessing pairwise contribution of causes to mortality. Cluster analysis techniques and weighting multiple causes were found to be newly emerging applications, applied only since 2016 and used in less than 3% of articles. Irrespective of the methodological approach, all the included articles demonstrated that multiple cause analysis complements the single UC approach by increasing the descriptive utility of the data and improving the quantification of causal attribution to mortality at both individual and population level.

Improved data quality and access to population level mortality data from vital registration systems may have facilitated increased use of multiple causes data over time.^74^ The review showed that for descriptive research questions, simple measures such as the numbers of deaths as well as death proportions and rates based on any-mention of a specific disease or condition may be sufficient in many cases. However, the complexity in structure of multiple causes data in terms of differentiating between causes listed in Part 1 and Part II of the death certificate, the application of rules for selection of the UC of death, and variations in death certification practices between countries and over time, pose challenges in interpreting analyses using multiple causes.^3,32,61,75,76^ The high frequency of analyses on drug-related mortality among the included articles likely arises as the ICD-10 coding mechanisms stipulate that the manner of death (intentional/unintentional poisoning) is routinely reported as the UC with the drug class represented by the associated causes; thus requiring analysis of the multiple causes.

Our audit of international experts coincided with a peak in the COVID-19 cases in Europe; the reliance on these public health experts during a period of heavy workload and uncertainty, may have contributed to the low response rate. Furthermore, analysis of multiple cause of death requires data from vital registrations, which are not available in all countries, thereby limiting the audit to high-income countries.

We could not identify a previous review capturing all contemporary methods used to assess multiple causes of death. Of the four narrative reviews identified from our search: two were published before 1990^10,16^ at which time there were 47 articles (81% based on US deaths data) and the other two^14,15^ were published over a decade ago and neither assessed the range of analytical methods that had been used. Only methods based on any-mention and pairwise assessment of causes were captured in the existing reviews.

The four groups of methods that we identified are distinct but complementary, and each has important practical applications. For instance, descriptive analyses based on any-mention provide useful contextual information to highlight the extent to which multiple causes play a role either as part of a causal sequence or through association between various causes listed on the death certificate. More specifically, while the SRMU is a descriptive measure, it reveals the extent of the potential contribution to mortality of causes that are not deemed to be the UC.^13,32,77^

However, changes in coding practices over time may influence the magnitude of underlying cause mortality from death certificates. This was observed in the case of diabetes^4^ and other conditions^29,78,79^ from the transition between the 9^th^ and 10^th^ revisions on the ICD; hence analyses of ‘any-mention’ rates might offer a better perspective of mortality trends, than rates based on the UC. While cause-related rates based on any-mention are simple to apply, they can be misleading in the context of overall mortality as each death is counted as many times as there are mentions.^69^

Assessing pairwise contribution to mortality is advantageous when investigating the relationships between two causes of death. Unlike basic summary statistics that consider one cause at a time, these methods consider two causes simultaneously; they were used frequently to evaluate external cause epidemiology, specifically to assess the nature of poisoning and injury related to exposure to drugs and alcohol.^80-84^ While the nature of the coding process mandates the use of multiple causes of death for assessing injuries sustained from external causes, descriptive pairwise analysis was regularly applied to infectious diseases^85-89^ and chronic diseases.^3,90-96^

More complex applications such as the assessment of mortality odds ratios were applied to assess the strength of relationships between disease on the death certificates. (e.g.^49,59,97-100^ H the disease associations based on deaths data alone do not imply causation; they evaluate whether the presence of a particular cause increases or decreases the probability of death from another cause,^14,47^ and statistical tests pertaining to strength of association between the causes should be interpreted with some caution^13^ as statistical assumptions, for example, of independence of causes, may not be valid because both causes contributed to death. Indeed, the associations between several causes mentioned on the death certificates are more frequent than would be expected by a random occurrence of the causes.^13^ Further, nearly all estimates of the associations between causes mentioned on the death certificates may be influenced by some level of Berkson’s paradox or collider bias.^101,102^ This is a form of selection bias that occurs when both the exposure and outcome variables (the two causes of death) influence the inclusion of participants in a study (death certificate data).^101^

Methods for grouping causes of death facilitate assessment of the complex relationships between causes that may go unnoticed by pairwise analysis. The pattern of diseases and risks leading to death may not be homogenous for the whole population, and methods that cluster deaths according to patterns in contributing causes can identify groups of individuals with specific combination of causes of death providing additional insight for setting targeted preventative interventions.^64^ A challenge with cluster analysis methods for grouping causes of death is that they are exploratory in nature and different clustering algorithms may result in different outcomes.^103-105^ Being data driven, the characteristics of the clusters are limited to the data, making it difficult to generalise the findings to populations in other settings. Furthermore, applications such as social network analysis describe the ‘closeness’ of causes of death, but further quantification of cause-related mortality is not possible.

On the other hand, user-defined groupings of causes, for example, those based on known disease-risk associations, are especially useful for examining the joint contribution and impact on mortality trends^61^ that cannot be captured by pairwise comparison.

The release of the ICD-11 for current implementation worldwide offers additional flexibility designed to enhance the evidence for informing better health systems.^106,107^ Of specific significance is the ‘post-coordination’ feature which allows combining specific codes into a cluster of relevant clinical attributes. While specific applications have not yet been defined for mortality, future application of multiple cause methods that group diseases based on multiple cause patterns or according to known disease-risk associations, could provide evidence to inform meaningful clusters for application in ICD-11.

Multiple-cause weighting methods have been developed to facilitate the measurement of overall magnitude of the contribution of a specific cause to population levels of mortality.^1,69^ A major advantage of these methods is the preservation of the counting unit (deaths) enabling derivation of a broad range of mortality indicators based on the weighted counts of each cause (e.g. age-standardised rates, years of life lost). This method overcomes the limitation of double-counting deaths for as many causes as present when using any-mention approaches.^69^ While the arbitrary nature of weights in MC-weighting strategies poses a limitation, the incorporation of methods for considering causal pathways of diseases by weighting only the UC and causes in Part II of the death certificate, and application of multiple cause weighting methods require careful consideration of the weighting strategy, the cause list and the handling of ill-defined causes,^73^ these methods offer a richer perspective for population health monitoring.^69^

The choice of the methods for analysing MC is dependent on the research question of interest. Irrespective of the methodological approach, MC analysis complements the single UC approach, uses useful information that is usually ignored and offers an additional perspective of the causes that contribute to death. Though there are distinct limitations around individual multiple cause methods, the broad range of methods described here offer a toolkit, which in combination can offer a richer perspective for population health monitoring and policy development.

To the best of our knowledge, this study is the largest review to date, comprehensively capturing statistical methods used for analysing multiple cause of death data, including many papers (>430) with two independent reviewers supplemented by an audit of international experts. A further strength is the use of a systematic approach to identify relevant studies, considering papers published in multiple languages. The inclusion of articles from a range of years, countries and languages revealed the breadth and diversity of applications of MC analysis.

This review is atypical of systematic reviews in that non-significant results and publication bias were not relevant. Publications largely represent data from countries with national vital statistics collections.

The very specific methods are not always apparent in the articles included here. For example, there is often little transparency around how duplicate mentions of causes and ill-defined causes are handled. As well, the terminologies used can be inconsistent (for example, contributing causes is often used to refer to non-UCs, but can also have a specific meaning referring to the causes reported in Part II of the death certificate). Additionally, factors that are known to affect multiple causes statistics such as the size and structure of the deaths certificate^13,24,60^ are not always apparent.

Our review showed that infectious diseases commonly assessed using multiple cause methods, for example, to ascertain socio-demographic differentials, to identify associated health conditions, and to assess the impact of health interventions to inform targeted prevention strategies.^71,86,108,109^ With new and emerging infectious diseases, MC data is crucial for descriptive epidemiology and for providing evidence to inform prevention strategies. Since declaration of COVID-19 as a pandemic in March 2020, the WHO implemented rules for ascertaining when a death was due to COVID-19 (i.e. the UC) noting that under certain circumstances COVID-19 should be recorded somewhere on the death certificate.^110^ While we identified only one COVID-related article (due to the timing of our database search), MC methods were since applied to ascertain associations between contributing conditions and complications, assess changes in the pathological patterns, and identify significant sociodemographic variation in COVID-related deaths,^111-114^ leading in some circumstances to improved survival.^115^ Importantly, the recent emergence of COVID-19 as a leading UC^116^ may significantly alter proportional mortality from other UCs. As such future analyses of multiple causes will be necessary for monitoring trends in COVID-related mortality as well as the impact of COVID-19 on other causes of death. As well, future sensitivity analyses that assess the impact of removing the non-underlying condition (by varying the weight ascribed to the UC)^69^ may facilitate assessment of competing causes of death where COVID-19 has become a major UC.

## Conclusion

The results from this review confirm that international interest is ongoing and increasing. This review provides the most comprehensive overview of multiple cause analytical methods and practices. The diversity of methods offers a toolkit for the analysis of these data which are becomingly increasingly important for understanding the complex involvement of multiple diseases in causing death across a range of settings including surveillance, policy, planning and research.

## Data Availability

The review included publicly available original research articles. All data produced in expert consultations are available upon reasonable request to the authors.

## References

1. Piffaretti C, Moreno-Betancur M, Lamarche-Vadel A, Rey G. Quantifying cause-related mortality by weighting multiple causes of death. Bull World Health Organ 2016;94(12):870–879.

2. Rao C. Medical certification of cause of death for COVID-19. Bull World Health Organ 2020;98(5):298–298a.

3. Desesquelles A, Demuru E, Salvatore MA, Pappagallo M, Frova L, Mesle F, Egidi V. Mortality from Alzheimer’s disease, Parkinson’s disease, and dementias in France and Italy: a comparison using the multiple cause-of-death approach. J Aging Health 2014;26(2):283–315.

4. Morrell S, Taylor R, Nand D, Rao C. Changes in proportional mortality from diabetes and circulatory disease in Mauritius and Fiji: possible effects of coding and certification. BMC Public Health 2019;19(1):481.

5. Nguyen-Nielsen M, Moller H, Tjonneland A, Borre M. Causes of death in men with prostate cancer: Results from the Danish Prostate Cancer Registry (DAPROCAdata). Cancer Epidemiol 2019;59:249–257.

6. Pinault L, Brauer M, Crouse DL, Weichenthal S, Erickson A, van Donkelaar A, Martin RV, Charbonneau S, Hystad P, Brook JR, Tjepkema M, Christidis T, Menard R, Robichaud A, Burnett RT. Diabetes status and susceptibility to the effects of PM2.5 exposure on cardiovascular mortality in a national canadian cohort. Epidemiology 2018;29(6):784–794.

7. D’Amico M, Agozzino E, Biagino A, Simonetti A, Marinelli P. Ill-defined and multiple causes on death certificates--a study of misclassification in mortality statistics. Eur J Epidemiol 1999;15(2):141–8.

8. Mannino DM, Brown C, Giovino GA. Obstructive lung disease deaths in the United States from 1979 through 1993. An analysis using multiple-cause mortality data. Am J Respir Crit Care Med 1997;156(3 Pt 1):814–8.

9. World Health Organization. International statistical classification of diseases and related health problems. - 10th revision, Fifth edition. Vol. 2. Geneva: World Health Organization, 2016.

10. Israel RA, Rosenberg HM, Curtin LR. Analytical potential for multiple cause-of-death data. [Review] [59 refs]. Am J Epidemiol. 124(2):161–79, 1986 Aug. 1986.

11. Stallard E. Underlying and multiple case mortality advanced ages: United States 1980-1998. North American Actuarial Journal 2002;6(3):64–87.

12. Chamblee RF, Evans MC. New dimensions in cause of death statistics. Am J Public Health 1982;72(11):1265–70.

13. Desesquelles A, Salvatore MA, Frova L, Pace M, Pappagallo M, Mesle F, Egidi V. Revisiting the mortality of France and Italy with the multiple-cause-of-death approach. Demogr Res 2010;23:771–805.

14. Bah S. Using multiple-cause mortality data to resolve conflicting information on trends in maternal mortality in South Africa. S Afr Med J 2006;96(4):308.

15. Bah S, Rahman MM. On measures of association for multiplecause mortality: Do we need more measures? Canadian Studies in Population 2011;38(3-4):93–104.

16. Puffer RR. New approaches for epidemiologic studies of mortality statistics. Bull Pan Am Health Organ 1989;23(4):365–83.

17. Bah S. Measures of multiple-cause mortality: a synthesis and a notational framework. Genus 2009;65(2):29–43.

18. Paik JM, Golabi P, Biswas R, Alqahtani S, Venkatesan C, Younossi ZM. Nonalcoholic fatty liver disease and alcoholic liver disease are major drivers of liver mortality in the United States. Hepatol Commun 2020;4(6):890–903.

19. Rodriguez F, Blum MR, Falasinnu T, Hastings KG, Hu J, Cullen MR, Palaniappan LP. Diabetes-attributable mortality in the United States from 2003 to 2016 using a multiple-cause-of-death approach. Diabetes Res Clin Pract 2019;148:169–178.

20. Simmons R, Ireland G, Ijaz S, Ramsay M, Mandal S. Causes of death among persons diagnosed with hepatitis C infection in the pre- and post-DAA era in England: A record linkage study. J Viral Hepat 2019;26(7):873–880.

21. Avouac J, Amrouche F, Meune C, Rey G, Kahan A, Allanore Y. Mortality profile of patients with rheumatoid arthritis in France and its change in 10 years. Semin Arthritis Rheum 2017;46(5):537–543.

22. Park CB, Yokoyama E, Tokuyama GH. Medical conditions at death among the caucasian and Japanese elderly in Hawaii - analysis of multiple causes of death, 1976-78. Journal of Clinical Epidemiology 1991;44(6):519–530.

23. Garcia Benavides F, Godoy C, Perez S, Bolumar F. Codificación Múltiple de las Causas de Muerte: de Morir "Por" a Morir "Con". Gac Sanit 1992;6(29):53–57.

24. Wall MM, Huang J, Oswald J, McCullen D. Factors associated with reporting multiple causes of death. BMC Med Res Methodol 2005;5(1):4.

25. Zargar AH, Wani AI, Masoodi SR, Bashir MI, Laway BA, Gupta VK, Wani FA. Causes of mortality in diabetes mellitus: data from a tertiary teaching hospital in India. Postgrad Med J 2009;85(1003):227–32.

26. Nembhard WN, Pathak EB, Schocken DD. Racial/ethnic disparities in mortality related to congenital heart defects among children and adults in the United States. Ethn Dis 2008;18(4):442–9.

27. Thomas SL, Griffiths C, Smeeth L, Rooney C, Hall AJ. Burden of mortality associated with autoimmune diseases among females in the United Kingdom. Am J Public Health 2010;100(11):2279–87.

28. Goldacre MJ, Duncan M, Griffith M, Turner MR. Trends in death certification for multiple sclerosis, motor neuron disease, Parkinson’s disease and epilepsy in English populations 1979-2006. J Neurol 2010;257(5):706–15.

29. Goldacre MJ, Duncan ME, Griffith M, Cook-Mozaffari P. Psychiatric disorders certified on death certificates in an English population. Soc Psychiatry Psychiatr Epidemiol 2006;41(5):409–14.

30. Jansson B, Johansson LA, Rosen M, Svanstrom L. National adaptations of the ICD rules for classification--a problem in the evaluation of cause-of-death trends. Journal of Clinical Epidemiology. 50(4):367–75, 1997 Apr. 1997.

31. Lu TH, Hsiao A, Chang PC, Chao YC, Hsu CC, Peng HC, Chen LH, Kawachi I. Counting injury deaths: a comparison of two definitions and two countries. Inj Prev 2013;21(e1):e127–32.

32. Goldberger N, Applbaum Y, Meron J, Haklai Z. High Israeli mortality rates from diabetes and renal failure - Can international comparison of multiple causes of death reflect differences in choice of underlying cause? Isr J Health Policy Res 2015;4:31.

33. Désesquelles A, Gamboni A, Demuru E, Barbieri M, Denissov G, Egidi V, Frova L, Pappagallo M, Goldberger N, Grundy E, Marshall C, Meslé F, Pechholdova M, Sakkeus L. We only die once… but from how many causes? Population and Societies 2016;534(1):1–4.

34. Désesquelles AD E.; Mésle, F.; Salvatore, M A. Cause-specific mortality analysis: Is the underlying cause of death sufficient? Revue Quetelet 2014.

35. Cummings PL, Kuo T, Javanbakht M, Sorvillo F. Trends, productivity losses, and associated medical conditions among toxoplasmosis deaths in the United States, 2000-2010. Am J Trop Med Hyg 2014;91(5):959–64.

36. Manton KG, Stallard E, Poss SS. Estimates of U.S. multiple cause life tables. Demography 1980;17(1):85–102.

37. Manton KG, Stallard E. Temporal trends in U. S. multiple cause of death mortality data: 1968 to 1977. Demography 1982;19(4):527–47.

38. Manton KGB, H. M. CVD mortality, 1968-1978: observations and implications. Stroke 1984.

39. Manton KG. Cause specific mortality patterns among the oldest old: multiple cause of death trends 1968 to 1980. J Gerontol 1986;41(2):282–9.

40. Moussa MA, El Sayed AM, Sugathan TN, Khogali MM, Verma D. Analysis of underlying and multiple-cause mortality data. Genus 1992;48(1-2):89–105.

41. Makela P. Alcohol-related mortality by age and sex and its impact on life expectancy - Estimates based on the Finnish death register. Eur J Public Health 1998;8(1):43–51.

42. Li SQ, Cunningham J, Cass A. Renal-related deaths in Australia 1997-1999. Intern Med J 2004;34(5):259–65.

43. Yoon YH, Stinson FS, Yi HY, Dufour MC. Accidental alcohol poisoning mortality in the United States, 1996-1998. Alcohol Res Health 2003;27(1):110–8.

44. Mannino DM, Ford E, Giovino GA, Thun M. Lung cancer deaths in the United States from 1979 to 1992: an analysis using multiple-cause mortality data. Int J Epidemiol 1998;27(2):159–66.

45. Chamblee RF, Evans MC, Patten DG, Pearce JS. Injuries causing death: Their nature, external causes, and associated diseases. J Safety Res 1983;14(1):21–35.

46. McCoy L, Redelings M, Sorvillo F, Simon P. A multiple cause-of-death analysis of asthma mortality in the United States, 1990-2001. J Asthma 2005;42(9):757–63.

47. Redelings MD, Wise M, Sorvillo F. Using multiple cause-of-death data to investigate associations and causality between conditions listed on the death certificate. Am J Epidemiol 2007;166(1):104–8.

48. Wickramasekaran RN, Sorvillo F, Kuo T. Legionnaires’ disease and associated comorbid conditions as causes of death in the U.S., 2000-2010. Public Health Rep 2015;130(3):222–9.

49. Wilkins K, Parsons GF, Gentleman JF, Forbes WF. Deaths due to dementia: An analysis of multiple-cause-of-death data. Chronic Dis Can 1999;20(1):26–35.

50. Arif N, Yousfi S, Vinnard C. Deaths from necrotizing fasciitis in the United States, 2003-2013. Epidemiol Infect 2016;144(6):1338–44.

51. Barragan NC, Moschetti K, Smith LV, Sorvillo F, Kuo T. Differential declines in syphilis-related mortality in the United States, 2000-2014. American Journal of Infection Control 2017;45(4):417–420.

52. Croker C, Reporter R, Redelings M, Mascola L. Strongyloidiasis-related deaths in the United States, 1991-2006. Am J Trop Med Hyg 2010;83(2):422–426.

53. Cummings PL, Sorvillo F, Kuo T. Salmonellosis-related mortality in the United States, 1990-2006. Foodborne Pathog Dis 2010;7(11):1393–9.

54. Barragan NC, Sorvillo F, Kuo T. Cryptococcosis-related deaths and associated medical conditions in the United States, 2000-2010. Mycoses 2014;57(12):741–6.

55. Rockett IR, Lian Y, Stack S, Ducatman AM, Wang S. Discrepant comorbidity between minority and white suicides: a national multiple cause-of-death analysis. BMC Psychiatry 2009;9:10.

56. Rockett IR, Wang S, Lian Y, Stack S. Suicide-associated comorbidity among US males and females: a multiple cause-of-death analysis. Inj Prev 2007;13(5):311–5.

57. Vinnard C, Longworth S, Mezochow A, Patrawalla A, Kreiswirth BN, Hamilton K. Deaths related to nontuberculous mycobacterial infections in the United States, 1999-2014. Ann Am Thorac Soc 2016;13(11):1951–1955.

58. Whiteside YO, Selik R, An Q, Huang T, Karch D, Hernandez AL, Hall HI. Comparison of rates of death having any death-certificate mention of heart, kidney, or liver disease among persons diagnosed with HIV infection with those in the general US population, 2009-2011. Open AIDS J 2015;9:14–22.

59. Fedeli U, Avossa F, Guzzinati S, Bovo E, Saugo M. Trends in mortality from chronic liver disease. Ann Epidemiol 2014;24(7):522–6.

60. Desesquelles AF, Salvatore MA, Pappagallo M, Frova L, Pace M, Mesle F, Egidi V. Analysing multiple causes of death: which methods for which data? An application to the cancer-related mortality in France and Italy. European Journal of Population-Revue Europeenne De Demographie 2012;28(4):467–498.

61. Adair T, Lopez AD. The role of overweight and obesity in adverse cardiovascular disease mortality trends: an analysis of multiple cause of death data from Australia and the USA. BMC Med 2020;18(1):199.

62. Yoon YH, Chen CM, Yi HY, Moss HB. Effect of comorbid alcohol and drug use disorders on premature death among unipolar and bipolar disorder decedents in the United States, 1999 to 2006. Compr Psychiatry 2011;52(5):453–64.

63. Villela PB, Klein CH, Oliveira GMM. Cerebrovascular and hypertensive diseases as multiple causes of death in Brazil from 2004 to 2013. Public Health 2018;161:36–42.

64. Barbieri M, Desesquelles A, Egidi V, Demuru E, Frova L, Mesle F, Pappagallo M. Obesity-related mortality in France, Italy, and the United States: a comparison using multiple cause-of-death analysis. Int J Public Health 2017;62(6):623–629.

65. Alexander MJ, Kiang MV, Barbieri M. Trends in black and white opioid mortality in the United States, 1979-2015. Epidemiology 2018;29(5):707–715.

66. Egidi V, Salvatore MA, Rivellini G, D’Angelo S. A network approach to studying cause-of-death interrelations. Demogr Res 2018;38(1):373–400.

67. Jiang H, Wu H, Wang MD. Causes of death in the United States, 1999 to 2014. 2017 IEEE EMBS International Conference on Biomedical and Health Informatics, BHI 2017 2017:177–180.

68. Hassanzadeh HR, Sha Y, Wang MD, Ieee. DeepDeath: Learning to predict the underlying cause of death with big data. 2017 39th Annual International Conference of the Ieee Engineering in Medicine and Biology Society 2017:3373–3376.

69. Moreno-Betancur M, Sadaoui H, Piffaretti C, Rey G. Survival analysis with multiple causes of death extending the competing risks model. Epidemiology 2017;28(1):12–19.

70. Gonzalez LF, Jo AHS, Garcia CAR. Weighted mortality method according to multiple causes of death. Finlay 2019;9(3):197–209.

71. Breger TL, Edwards JK, Cole SR, Saag M, Rebeiro PF, Moore RD, Eron JJ. Estimating a set of mortality risk functions with multiple contributing causes of death. Epidemiology 2020;31(5):704–712.

72. Xie SH, Chen H, Lagergren J. Causes of death in patients diagnosed with gastric adenocarcinoma in Sweden, 1970-2014: A population-based study. Cancer Sci 2020;111(7):2451–2459.

73. Bishop KA, Moreno-Betancur, M., Balogun, S., Eynstone-Hinkins, J., Moran, L., Rao, C., Banks, E., Korda, R.J., Gourley, M., Joshy, G. Quantifying cause-related mortality in Australia incorporating multiple causes: observed patterns, trends, and practical considerations. (in press) 2022.

74. Rao C, Adair T, Bain C, Doi SA. Mortality from diabetic renal disease: a hidden epidemic. Eur J Public Health 2011;22(2):280–4.

75. Lu TH, Hsu PY, Bjorkenstam C, Anderson RN. Certifying diabetes-related cause-of-death: a comparison of inappropriate certification statements in Sweden, Taiwan and the USA. Diabetologia 2006;49(12):2878–81.

76. Adair T, Rao C. Changes in certification of diabetes with cardiovascular diseases increased reported diabetes mortality in Australia and the United States. J Clin Epidemiol 2010;63(2):199–204.

77. Buschner A, Grunwald-Muhlberger A. Influence of methodological changes on unicausal cause-of-death statistics and potentials of a multicausal data basis. Bundesgesundheitsblatt-Gesundheitsforschung-Gesundheitsschutz 2019;62(12):1476–1484.

78. Goldacre MJ, Duncan ME, Griffith M. Death rates for asthma in English populations 1979-2007: comparison of underlying cause and all certified causes. Public Health 2012;126(5):386–93.

79. Jansson B, Ahmed N. Epilepsy and injury mortality in Sweden--the importance of changes in coding practice. Seizure 2002;11(6):361–70.

80. Chitty KM, Schumann JL, Moran LL, Chong DG, Hurzeler TP, Buckley NA. Reporting of alcohol as a contributor to death in Australian national suicide statistics and its relationship to post-mortem alcohol concentrations. Addiction 2021;116(3):506–513.

81. Kiadaliri AA, Petersson IF, Englund M. Educational inequalities in mortality associated with rheumatoid arthritis and other musculoskeletal disorders in Sweden. BMC Musculoskeletal Disorders 2019;20(1):83.

82. Kiadaliri AA, Rosengren BE, Englund M. Fracture-related mortality in southern Sweden: A multiple cause of death analysis, 1998-2014. Injury 2018;49(2):236–242.

83. Turner C, Chandrakumar D, Rowe C, Santos GM, Riley ED, Coffin PO. Cross-sectional cause of death comparisons for stimulant and opioid mortality in San Francisco, 2005-2015. Drug Alcohol Depend 2018;185:305–312.

84. Parks SE, Kegler SR, Annest JL, Mercy JA. Characteristics of fatal abusive head trauma among children in the USA: 2003-2007: an application of the CDC operational case definition to national vital statistics data. Inj Prev 2012;18(3):193–9.

85. Fedeli U, Grande E, Grippo F, Frova L. Mortality associated with hepatitis C and hepatitis B virus infection: A nationwide study on multiple causes of death data. World J Gastroenterol 2017;23(10):1866–1871.

86. Ford MM, Desai PS, Maduro G, Laraque F. Neighborhood inequalities in Hepatitis C mortality: Spatial and temporal patterns and associated factors. J Urban Health 2017;94(5):746–755.

87. Grande E, Zucchetto A, Suligoi B, Grippo F, Pappagallo M, Virdone S, Camoni L, Taborelli M, Regine V, Serraino D, Frova L. Multiple cause-of-death data among people with AIDS in Italy: a nationwide cross-sectional study. Popul Health Metr 2017;15(1):19.

88. Pechholdova M. Sepsis-related mortality in the Czech Republic: multiple causes of death analysis. Epidemiologie, Mikrobiologie, Imunologie. 66(2):73–79, Summer 2017. 2017.

89. da Silva LR, Araújo ETH, Carvalho ML, Almeida CAPL, da Silva Oliveira AD, de Carvalho PMG, Rodrigues TS, Campelo V. Epidemiological situation of acquired immunodeficiency syndrome (Aids)-related mortality in a municipality in northeastern brazil. a retrospective cross-sectional study. Sao Paulo Medical Journal 2018;136(1):37–43.

90. Duncan ME, Pitcher A, Goldacre MJ. Atrial fibrillation as a cause of death increased steeply in England between 1995 and 2010. Europace 2014;16(6):797–802.

91. Garcia-Ptacek S, Kareholt I, Cermakova P, Rizzuto D, Religa D, Eriksdotter M. Causes of death according to death certificates in individuals with dementia: A cohort from the Swedish dementia registry. J Am Geriatr Soc 2016;64(11):e137–e142.

92. Takamori A, Takahashi I, Kasagi F, Suyama A, Ozasa K, Yanagawa T. Mortality analysis of the Life Span Study (LSS) cohort taking into account multiple causes of death indicated in death certificates. Radiat Res 2017;187(1):20–31.

93. Seuc AH, Fernandez-Gonzalez L, Mirabal M. Comparative disease assessment: a multi-causal approach for estimating the burden of mortality. Journal of Public Health 2020;30(3):665–673.

94. Amaral TLM, Amaral CA, Miranda Filho AL, Monteiro GTR. Trends and multiple causes of death due to chronic renal failure in a municipality in the Brazilian Amazon. Cien Saude Colet 2018;23(11):3821–3828.

95. Fedeli U, Schievano E, Lisiero M, Avossa F, Mastrangelo G, Saugo M. Descriptive epidemiology of chronic liver disease in northeastern Italy: an analysis of multiple causes of death. Popul Health Metr 2013;11(1):20.

96. Fedeli U, Schievano E, Targher G, Bonora E, Corti MC, Zoppini G. Estimating the real burden of cardiovascular mortality in diabetes. Eur Rev Med Pharmacol Sci 2019;12(15):6700–6706.

97. Redelings MDL, N. E.; Sorvillo, F. Pressure ulcers: more lethal than we thought? Adv Skin Wound Care 2005.

98. Ramos AN, Jr., Matida LH, Hearst N, Heukelbach J. Mortality in Brazilian children with HIV/AIDS: the role of non-AIDS-related conditions after highly active antiretroviral therapy introduction. AIDS Patient Care STDS 2011;25(12):713–8.

99. Chang CY, Lu TH, Cheng TJ. Trends in reporting injury as a cause of death among people with epilepsy in the U.S., 1981-2010. Seizure 2014;23(10):836–43.

100. Grippo F, Pappagallo M, Burgio A, Crialesi R. Drug induced mortality: A multiple cause approach on italian causes of death register. Epidemiology Biostatistics and Public Health 2015;12:e–1.

101. Redelings MD, Wise M, Sorvillo F. Using multiple cause-of-death data to investigate associations and causality between conditions listed on the death certificate. American journal of epidemiology 2007;166(1):104–108.

102. Laanani M, Viallon V, Coste J, Rey G. Collider and Reporting Biases Involved in the Analyses of Cause of Death Associations in Death Certificates: an Illustration with Cancer and Suicide. Preprint from Research Square 2021:1–18.

103. Newcomer SR, Steiner JF, Bayliss EA. Identifying subgroups of complex patients with cluster analysis. The American journal of managed care 2011;17(8):e324–32.

104. Frova L, Salvatore MA, Pappagallo M, Egidi V. The multiple cause of death approach to analyse mortality patterns. Genus 2009;65(1):1–21.

105. Roso-Llorach A, Violán C, Foguet-Boreu Q, Rodriguez-Blanco T, Pons-Vigués M, Pujol-Ribera E, Valderas JM. Comparative analysis of methods for identifying multimorbidity patterns: a study of ‘real-world’data. BMJ open 2018;8(3):e018986.

106. Mabon K, Steinum O, Chute CG. Postcoordination of codes in ICD-11. BMC Med Inform Decis Mak 2022;21(Suppl 6):379.

107. World Health Organization. International Classification of Diseases Eleventh Revision (ICD-11). Geneva: World Health Organization, 2022.

108. Domingues CSB, Waldman EA. Causes of death among people living with AIDS in the Pre- and Post-HAART Eras in the city of S(a)over-tildeo Paulo, Brazil. PLoS One 2014;9(12).

109. Goldstein E, Lipsitch M. The relation between prescribing of different antibiotics and rates of mortality with sepsis in US adults. BMC Infect Dis 2020;20(1):169.

110. World Health Organization. International Guidleines for Certification and Classification (Coding) of COVID-19 as Cause of Death Geneva: World Health Organization, 2020.

111. Grippo F, Navarra S, Orsi C, Manno V, Grande E, Crialesi R, Frova L, Marchetti S, Pappagallo M, Simeoni S, Di Pasquale L, Carinci A, Donfrancesco C, Lo Noce C, Palmieri L, Onder G, Minelli G, Italian National Institute of Health Covid-Mortality G. The role of COVID-19 in the death of SARS-CoV-2-positive patients: a study based on death certificates. J Clin Med 2020;9(11).

112. Christensen RAG, Arneja J, St Cyr K, Sturrock SL, Brooks JD. The association of estimated cardiorespiratory fitness with COVID-19 incidence and mortality: A cohort study. PLoS One 2021;16(5):e0250508.

113. Nogales Vasconcelos AM, Ishitani L, Abreu DMX, Franca E. Covid adult mortality in Brazil: an analysis of multiple causes of death. Front Public Health 2021;9:788932.

114. Woodward M, Peters SAE, Harris K. Social deprivation as a risk factor for COVID-19 mortality among women and men in the UK Biobank: nature of risk and context suggests that social interventions are essential to mitigate the effects of future pandemics. J Epidemiol Community Health 2021;75(11):1050–1055.

115. Grippo F, Grande E, Maraschini A, Navarra S, Pappagallo M, Marchetti S, Crialesi R, Frova L, Orsi C, Simeoni S, Carinci A, Loreto G, Donfrancesco C, Lo Noce C, Palmieri L, Andrianou X, Urdiales AM, Onder G, Minelli G, Italian National Institute of Health C-MG. Evolution of pathology patterns in persons who died from COVID-19 in Italy: a national study based on death certificates. Front Med (Lausanne) 2021;8:645543.

116. Woolf SH, Chapman DA, Lee JH. COVID-19 as the leading cause of death in the United States. JAMA 2021;325(2):123–124.

