## Supplementary File 1 for "Analysis of multiple causes of death: a review of methods and practices"

SUPPLEMENTARY FILE 1 – SEARCH STRATEGY

**Analysis of multiple causes of death: a review of methods and practices**

**Table S1.1. Search strategy for different databases**

| **Database** | **Search strategy** |
| --- | --- |
| Scopus | ( ( "multiple cause* of" W/1 ( death OR mortality ) ) OR ( multiple W/3 "cause* of mortality" ) OR ( multiple W/3 "cause* of death" ) OR ( "multiple cause mortality" ) OR ( "contributory cause* of" W/1 ( death OR mortality ) ) OR ( "other cause* of" W/1 ( death OR mortality ) ) OR ( "underlying and contributory" W/3 ( death OR mortality ) ) AND ( "death certificate" OR "death record" OR "ICD code" OR "Cause of Death ICD Codes" ) ) OR ( "associated cause* of death" ) |
| Web of Science | TS = ("multiple cause* of death") OR TS=("multiple cause mortality") OR TS=("contributory cause* of death") OR TS=("contributing cause* of death") OR TS = ("underlying cause* of death") OR TS = ("associated cause* of death") |
| Medline | (multiple cause of death or multiple causes of death or multiple cause mortality or underlying cause of death or underlying causes of death or contributory cause of death or contributory causes of death or contributing cause of death or associated cause of death or associated causes of death).mp. |
| Pubmed | "multiple cause of death"[All Fields] OR "multiple causes of death"[All Fields] OR "multiple cause mortality"[All Fields] OR "contributory cause of death"[All Fields] OR "contributing cause of death"[All Fields] OR "underlying cause of death"[All Fields] OR "associated cause of death"[All Fields] |
