## Supplementary File 2 for "Analysis of multiple causes of death: a review of methods and practices"

SUPPLEMENTARY FILE 2 – REFERENCES FOR INCLUDED TEXTS

**Analysis of multiple causes of death: a review of methods and practices**

This supplement contains the references for articles included in the full text review. Table S2.1 indicates the reference numbers for articles identified as applying one of the assessed multiple cause methods.

**Table S2.1.** References for included texts by methods category

| **Methods category (number of articles)** | **Reference numbers for included texts** |
| --- | --- |
| Any mention method (n=377) | 1-377 |
| Pairwise assessment method (n=245) | 1–191, 378–431 |
| Grouped causes method (n=9) | 55, 162, 173, 188, 192, 196, 219, 433, 434 |
| MC weighting method (n=4) | 246, 306, 322, 371, 432 |

**References for included texts**

1. Adekoya N, Majumder R. Fatal traumatic brain injury, West Virginia, 1989-1998. Public Health Rep 2004;119(5):486-92.
2. Adih WK, Selik RM, Hu X. Trends in Diseases Reported on US Death Certificates That Mentioned HIV Infection, 1996-2006. J Int Assoc Physicians AIDS Care (Chic) 2011;10(1):5-11.
3. Alicandro G, Frova L, Di Fraia G, Colombo C. Cystic fibrosis mortality trend in Italy from 1970 to 2011. J Cyst Fibros 2015;14(2):267-74.
4. Amaral TLM, Amaral CA, Miranda Filho AL, Monteiro GTR. Trends and multiple causes of death due to chronic renal failure in a municipality in the Brazilian Amazon. Cien Saude Colet 2018;23(11):3821-3828.
5. Andreasyan K, Hoy WE. Renal-related deaths in Indigenous people in Queensland, Australia. Nephrology 2007;12(5):514-519.
6. Arif N, Yousfi S, Vinnard C. Deaths from necrotizing fasciitis in the United States, 2003-2013. Epidemiol Infect 2016;144(6):1338-44.
7. Ascoli V, Minelli G, Kanieff M, et al. Cause-specific mortality in classic Kaposi's sarcoma: a population-based study in Italy (1995-2002). Br J Cancer 2009;101(7):1085-90.
8. Ascoli V, Minelli G, Kanieff M, Frova L, Conti S. Merkel cell carcinoma: a population-based study on mortality and the association with other cancers. Cancer Causes Control 2011;22(11):1521-7.
9. Avouac J, Amrouche F, Meune C, et al. Mortality profile of patients with rheumatoid arthritis in France and its change in 10 years. Semin Arthritis Rheum 2017;46(5):537-543.
10. Barragan NC, Sorvillo F, Kuo T. Cryptococcosis-related deaths and associated medical conditions in the United States, 2000-2010. Mycoses 2014;57(12):741-6.
11. Bate J, Ladhani S, Sharland M, et al. Infection-related mortality in children with malignancy in England and Wales, 2003-2005. Pediatr Blood Cancer 2009;53(3):371-4.
12. Baum HM, Manton KG. National trends in stroke-related mortality: a comparison of multiple cause mortality data with survey and other health data. Gerontologist 1987;27(3):293-300.
13. Benavides GF, Godoy C, Pérez S, Bolumar F. [Multiple codification of the causes of death: from dying "of" to dying "from"]. Gac Sanit 1992;6(29):53-7.
14. Bennion JR, Sorvillo F, Wise ME, Krishna S, Mascola L. Decreasing listeriosis mortality in the United States, 1990-2005. Clinical Infectious Diseases 2008;47(7):867-874.
15. Bi P, Parton KA, Whitby M. Co-existing conditions for deaths from infectious and parasitic diseases in Australia. Int J Infect Dis 2004;8(2):121-5.
16. Boslett AJ, Denham A, Hill EL. Using contributing causes of death improves prediction of opioid involvement in unclassified drug overdoses in US death records. Addiction 2020;115(7):1308-1317.
17. Boyle CA, Decoufle P, Holmgreen P. Contribution of developmental-disabilities to childhood mortality in the United-States - A multiple cause-of-death analysis. Am J Epidemiol 1994;138(8):665-665.
18. Cardoso BB, Kale PL. Coding pulmonary sepsis and mortality statistics in Rio de Janeiro, RJ. Rev Bras Epidemiol 2016;19(3):609-620.
19. Chamblee RF, Evans MC. New dimensions in cause of death statistics. Am J Public Health 1982;72(11):1265-70.
20. Chazal T, Lhote R, Rey G, et al. Giant-cell arteritis-related mortality in France: A multiple-cause-of-death analysis. Autoimmun Rev 2018;17(12):1219-1224.
21. Cheng Y, Han X, Luo Y, Xu W. Deaths of obstructive lung disease in the Yangpu district of Shanghai from 2003 through 2011: a multiple cause analysis. Chin Med J (Engl) 2014;127(9):1619-25.
22. Chiavegatto Filho ADP, Laurenti R, Gotlieb SLD, Jorge MHPDM. Malnutrition as an underlying or associated cause of death: Analysis of the quality of information on women of reproductive age. Revista Brasileira de Epidemiologia 2007;10(1):30-38.
23. Chorba TL, Holman RC, Clarke MJ, Evatt BL. Effects of HIV infection on age and cause of death for persons with hemophilia A in the United States. Am J Hematol 2001;66(4):229-40.
24. Coren S, Hewitt PL. Is anorexia nervosa associated with elevated rates of suicide? Am J Public Health 1998;88(8):1206-1207.
25. Costi LR, Iwamoto HM, Neves DCD, Caldas CAM. Mortality from systemic erythematosus lupus in Brazil: evaluation of causes according to the government health database. Rev Bras Reumatol Engl Ed 2017;57(6):574-582.
26. Craig Hooper W, Holman RC, Clarke MJ, Chorba TL. Trends in non-Hodgkin lymphoma (NHL) and HIV-associated NHL deaths in the United States. Am J Hematol 2001;66(3):159-166.
27. Crews DE, Stamler J, Dyer A. Conditions other than underlying cause of death listed on death certificates provide additional useful information for epidemiologic research. Epidemiology 1991;2(4):271-5.
28. Croker C, Reporter R, Redelings M, Mascola L. Strongyloidiasis-related deaths in the United States, 1991-2006. Am J Trop Med Hyg 2010;83(2):422-426.
29. Cummings PL, Sorvillo F, Kuo T. Salmonellosis-related mortality in the United States, 1990-2006. Foodborne Pathog Dis 2010;7(11):1393-9.
30. Cummings PL, Kuo T, Javanbakht M, Sorvillo F. Trends, productivity losses, and associated medical conditions among toxoplasmosis deaths in the United States, 2000-2010. Am J Trop Med Hyg 2014;91(5):959-64.
31. Cutter JL. Diabetes mortality in Singapore, 1991--a preliminary study. Singapore Med J 1998;39(7):311-8.
32. da Silva LR, Araújo ETH, Carvalho ML, et al. Epidemiological situation of acquired immunodeficiency syndrome (Aids)-related mortality in a municipality in northeastern brazil. a retrospective cross-sectional study. Sao Paulo Medical Journal 2018;136(1):37-43.
33. DeGiorgio CM, Curtis A, Carapetian A, et al. Why are epilepsy mortality rates rising in the United States? A population-based multiple cause-of-death study. BMJ Open 2020;10(8):e035767.
34. Désesquelles A, Meslé F. Intérêt de l’analyse des causes multiples dans l’étude de la mortalité aux grands âges : l’exemple français. Cahiers québécois de démographie 2005;33(1):83-116.
35. Désesquelles A, Salvatore MA, Frova L, et al. Revisiting the mortality of France and Italy with the multiple-cause-of-death approach. Demogr Res 2010;23:771-805.
36. Désesquelles AF, Salvatore MA, Pappagallo M, et al. Analysing multiple causes of death: Which ethods for which data? An application to the cancer-related mortality in France and Italy. European Journal of Population-Revue Europeenne De Demographie 2012;28(4):467-498.
37. Désesquelles A, Demuru E, Salvatore MA, et al. Mortality from Alzheimer's disease, Parkinson's disease, and dementias in France and Italy: a comparison using the multiple cause-of-death approach. J Aging Health 2014;26(2):283-315.
38. Désesquelles A, Demuru E, Pappagallo M, et al. After the epidemiologic transition: a reassessment of mortality from infectious diseases among over-65s in France and Italy. Int J Public Health 2015;60(8):961-7.
39. Dong C, Yoon YH, Chen CM, Hsiao-Ye Y. Heavy alcohol use and premature death from hepatocellular carcinoma in the United States, 1999-2006. J Stud Alcohol Drugs 2011;72(6):892-902.
40. Ducci RD, Cirino RH, de Oliveira RA, et al. Analysis of 621 death certificates issued from 1998 to 2007 in Curitiba, Brazil, mentioning epilepsy, epileptic seizures and/or status epilepticus. Seizure 2011;20(5):406-8.
41. Duncan ME, Goldacre MJ. Mortality trends for benign prostatic hyperplasia and prostate cancer in English populations 1979-2006. BJU Int 2011;107(1):40-5.
42. Duncan ME, Pitcher A, Goldacre MJ. Atrial fibrillation as a cause of death increased steeply in England between 1995 and 2010. Europace 2014;16(6):797-802.
43. Durkin A, Connolly S, O'Reilly D. Quantifying alcohol-related mortality: should alcohol-related contributory causes of death be included? Alcohol Alcohol 2010;45(4):374-8.
44. Ecclestone TC, Yeates DG, Goldacre MJ. Fall in population-based mortality from coronary heart disease negated in people with diabetes mellitus: data from England. Diabet Med 2015;32(10):1329-34.
45. Evans JM, Barnett KN, McMurdo ME, Morris AD. Reporting of diabetes on death certificates of 1872 people with type 2 diabetes in Tayside, Scotland. Eur J Public Health 2008;18(2):201-3.
46. Fearnley E, Li SQ, Guthridge S. Trends in chronic disease mortality in the Northern Territory Aboriginal population, 1997-2004: using underlying and multiple causes of death. Aust N Z J Public Health 2009;33(6):551-5.
47. Fedeli U, Avossa F, Goldoni CA, et al. Education level and chronic liver disease by aetiology: A proportional mortality study. Dig Liver Dis 2015;47(12):1082-5.
48. Fedeli U, Zoppini G, Goldoni CA, et al. Multiple causes of death analysis of chronic diseases: the example of diabetes. Popul Health Metr 2015;13:21.
49. Fedeli U, Piccinni P, Schievano E, Saugo M, Pellizzer G. Growing burden of sepsis-related mortality in northeastern Italy: a multiple causes of death analysis. BMC Infect Dis 2016;16:330.
50. Fedeli U, Grande E, Grippo F, Frova L. Mortality associated with hepatitis C and hepatitis B virus infection: A nationwide study on multiple causes of death data. World J Gastroenterol 2017;23(10):1866-1871.
51. Fedeli U, Schievano E. Increase in Parkinson's disease-related mortality among males in Northern Italy. Parkinsonism Relat Disord 2017;40:47-50.
52. Fedeli U, Schievano E, Targher G, et al. Estimating the real burden of cardiovascular mortality in diabetes. Eur Rev Med Pharmacol Sci 2019;12(15):6700-6706.
53. Fingerhut LA, Cox CS. Poisoning mortality, 1985-1995. Public Health Rep 1998;113(3):218-33.
54. Franco LJ, Mameri C, Pagliaro H, Iochida LC, Goldenberg P. Diabetes as underlying or associated cause of death in the State of S. Paulo, 1992. Rev Saude Publica 1998;32(3):237-245.
55. Frova L, Salvatore MA, Pappagallo M, Egidi V. The multiple cause of death approach to analyse mortality patterns. Genus 2009;65(1):1-21.
56. Garcia-Ptacek S, Kareholt I, Cermakova P, et al. Causes of death according to death certificates in individuals with dementia: A cohort from the Swedish Dementia Registry. J Am Geriatr Soc 2016;64(11):e137-e142.
57. Goldacre MJ, Mant D, Duncan M, Griffith M. Mortality from heart failure in an English population, 1979–2003: study of death certification. Journal of Epidemiology and Community Health 2005;59(9):782.
58. Goldacre MJ, Duncan ME, Griffith M, Davidson M. Trends in mortality from appendicitis and from gallstone disease in English populations, 1979-2006: study of multiple-cause coding of deaths. Postgrad Med J 2011;87(1026):245-50.
59. Goldacre MJ, Duncan ME. Death rates for acquired hypothyroidism and thyrotoxicosis in English populations (1979-2010): comparison of underlying cause and all certified causes. QJM2013;106(3):229-35.
60. Goldberger N, Applbaum Y, Meron J, Haklai Z. High Israeli mortality rates from diabetes and renal failure - Can international comparison of multiple causes of death reflect differences in choice of underlying cause? Isr J Health Policy Res 2015;4:31.
61. Gorina Y, Lentzner H. Multiple causes of death in old age. Aging Trends 2008(9):1-9.
62. Grande E, Zucchetto A, Suligoi B, et al. Multiple cause-of-death data among people with AIDS in Italy: a nationwide cross-sectional study. Popul Health Metr 2017;15(1):19.
63. Grande E, Grippo F, Frova L, et al. The increase of sepsis-related mortality in Italy: a nationwide study, 2003-2015. Eur J Clin Microbiol Infect Dis 2019;38(9):1701-1708.
64. Gravensteen IK, Ekeberg O, Thiblin I, et al. Psychoactive substances in natural and unnatural deaths in Norway and Sweden - a study on victims of suicide and accidents compared with natural deaths in psychiatric patients. BMC Psychiatry 2019;19(1):33.
65. Griffiths EC, Pedersen AB, Fenton A, Petchey OL. Reported co-infection deaths are more common in early adulthood and among similar infections. BMC Infectious Diseases 2015;15(1).
66. Grippo F, Pappagallo M, Burgio A, Crialesi R. Drug induced mortality: A multiple cause approach on Italian causes of death register. Epidemiology Biostatistics and Public Health 2015;12:e-1.
67. Grippo F, Désesquelles A, Pappagallo M, et al. Multi-morbidity and frailty at death: A new classification of death records for an ageing world. Popul Stud (Camb) 2020;74(3):437-449.
68. Gu K, Cowie CC, Harris MI. Mortality in adults with and without diabetes in a national cohort of the U.S. population, 1971-1993. Diabetes Care 1998;21(7):1138-45.
69. Guralnick L. Some problems in the use of multiple causes of death. J Chronic Dis 1966;19(9):979-90.
70. Harding AH, Darnton AJ. Asbestosis and mesothelioma among British asbestos workers (1971-2005). Am J Ind Med 2010;53(11):1070-80.
71. Harding K, Zhu F, Alotaibi M, et al. Multiple cause of death analysis in multiple sclerosis: A population-based study. Neurology 2020;94(8):e820-e829.
72. Harzke AJ, Baillargeon J, Paar DP, Pulvino J, Murray OJ. Chronic liver disease mortality among male prison inmates in Texas, 1989-2003. The American journal of gastroenterology 2009;104(6):1412-1419.
73. Horlander KR, Mannino DM, Leeper KV. Pulmonary embolism mortality in the United States, 1979-1998 - An analysis using multiple-cause mortality data. Arch Intern Med 2003;163(14):1711-1717.
74. Jaen G, Drowos J, Hennekens CH, Levine RS. Lower mortality rates from Cryptococcosis in women and whites with Human Immunodeficiency Virus in the United States. J Racial Ethn Health Disparities 2020;7(1):117-120.
75. Janssen TA. Importance of tabulating multiple causes of death. Am J Public Health Nations Health 1940;30(8):871-9.
76. Johnson NE, Christenson BA. Socio-demographic correlates of multiple causes of death: Real or artifactual? Population Research and Policy Review 1998;17(3):261-274.
77. Juel K, Helweg-Larsen K. Drug-related mortality in Denmark 1970-93. Scand J Public Health 1999;27(1):48-53.
78. Jung RS, Bennion JR, Sorvillo F, Bellomy A. Trends in tuberculosis mortality in the United States, 1990-2006: a population-based case-control study. Public Health Rep 2010;125(3):389-97.
79. Kandel DB, Hu MC, Griesler P, Wall M. Increases from 2002 to 2015 in prescription opioid overdose deaths in combination with other substances. Drug Alcohol Depend 2017;178:501-511.
80. Kenneson A, Kolor K, Yang Q, et al. Trends and racial disparities in muscular dystrophy deaths in the United States, 1983-1998: an analysis of multiple cause mortality data. Am J Med Genet A 2006;140(21):2289-97.
81. Kiadaliri AA, Turkiewicz A, Englund M. Mortality from Musculoskeletal Disorders including Rheumatoid Arthritis in Southern Sweden: A multiple-cause-of-death analysis, 1998-2014. J Rheumatol 2017;44(5):571-579.
82. Kiadaliri AA, Rosengren BE, Englund M. Fracture-related mortality in southern Sweden: A multiple cause of death analysis, 1998-2014. Injury 2018;49(2):236-242.
83. Kiadaliri AA, Rosengren BE, Englund M. Fall-related mortality in southern Sweden: a multiple cause of death analysis, 1998-2014. Inj Prev 2019;25(2):129-135.
84. Kishamawe C, Rumisha SF, Mremi IR, et al. Trends, patterns and causes of respiratory disease mortality among inpatients in Tanzania, 2006-2015. Trop Med Int Health 2019;24(1):91-100.
85. Kugeler KJ, Griffith KS, Gould LH, et al. A review of death certificates listing Lyme disease as a cause of death in the United States. Clin Infect Dis 2011;52(3):364-7.
86. Landes SD, Stevens JD, Turk MA. Obscuring effect of coding developmental disability as the underlying cause of death on mortality trends for adults with developmental disability: a cross-sectional study using US Mortality Data from 2012 to 2016. BMJ Open 2019;9(2):e026614.
87. Landes SD, Stevens JD, Turk MA. Cause of death in adults with Down syndrome in the United States. Disabil Health J 2020;13(4):100947.
88. Lee DS, Gona P, Albano I, et al. A systematic assessment of causes of death after heart failure onset in the ommunity impact of age at death, time period, and left ventricular systolic dysfunction. Circulation-Heart Failure 2011;4(1):36-U85.
89. Leone M, Chandra V, Schoenberg BS. Motor neuron disease in the United States, 1971 and 1973–1978: Patterns of mortality and associated conditions at the time of death. Neurology 1987;37(8):1339-1343.
90. Lethbridge L, Johnston GM, Turnbull G. Co-morbidities of persons dying of Parkinson's disease. Progress in Palliative Care 2013;21(3):140-145.
91. Li SQ, Cunningham J, Cass A. Renal-related deaths in Australia 1997-1999. Intern Med J 2004;34(5):259-65.
92. Lin JC, Nichol KL. Excess mortality due to pneumonia or influenza during influenza seasons among persons with acquired immunodeficiency syndrome. Arch Intern Med 2001;161(3):441-6.
93. Lott JP, Gross CP. Mortality from nonneoplastic skin disease in the United States. J Am Acad Dermatol 2014;70(1):47-54 e1.
94. Ly KN, Xing J, Klevens RM, et al. The increasing burden of mortality from viral hepatitis in the United States between 1999 and 2007. Ann Intern Med 2012;156(4):271-8.
95. Ly KN, Xing J, Klevens RM, Jiles RB, Holmberg SD. Causes of death and characteristics of decedents with viral hepatitis, United States, 2010. Clinical Infectious Diseases 2014;58(1):40-49.
96. Ly KN, Klevens RM. Trends in disease and complications of hepatitis A virus infection in the United States, 1999-2011: a new concern for adults. J Infect Dis 2015;212(2):176-82.
97. Mackenbach JP, Kunst AE, Lautenbach H, Oei YB, Bijlsma F. Competing causes of death: a death certificate study. J Clin Epidemiol 1997;50(10):1069-77.
98. Mackenbach JP, Kunst AE, Lautenbach H, Bijlsma F, Oei YB. Competing causes of death: an analysis using multiple-cause-of-death data from The Netherlands. Am J Epidemiol 1995;141(5):466-75.
99. Mannino DM, Ford E, Giovino GA, Thun M. Lung cancer deaths in the United States from 1979 to 1992: an analysis using multiple-cause mortality data. Int J Epidemiol 1998;27(2):159-66.
100. Masocco M, Kodra Y, Vichi M, et al. Mortality associated with neurofibromatosis type 1: a study based on Italian death certificates (1995-2006). Orphanet J Rare Dis 2011;6:11.
101. Maynard C, Lowy E, McDonell M, Fihn SD. Cause of death in Washington state veterans hospitalized with acute coronary syndromes in the veterans health administration. Popul Health Metr 2008;6:3.
102. McEwen LN, Kim C, Haan M, et al. Diabetes reporting as a cause of death: results from the Translating Research Into Action for Diabetes (TRIAD) study. Diabetes Care 2006;29(2):247-53.
103. McEwen LN, Karter AJ, Curb JD, et al. Temporal trends in recording of diabetes on death certificates: results from Translating Research Into Action for Diabetes (TRIAD). Diabetes Care 2011;34(7):1529-33.
104. McKenzie K, Chen L, Walker SM. Correlates of undefined cause of injury coded mortality data in Australia. Health Inf Manag 2009;38(1):8-14.
105. McNeil MM, Nash SL, Hajjeh RA, et al. Trends in mortality due to invasive mycotic diseases in the United States, 1980-1997. Clin Infect Dis 2001;33(5):641-7.
106. McPherson D, Griffiths C, Williams M, et al. Sepsis-associated mortality in England: an analysis of multiple cause of death data from 2001 to 2010. BMJ Open 2013;3(8).
107. Mendlein JM, Sattin RW, Waxweiler RJ, Lui KJ, McGee DL. Fall mortality and related medical conditions in the elderly: The association with pulmonary embolism. J Aging Health 1990;2(3):326-340.
108. Murray CJ, Dias RH, Kulkarni SC, et al. Improving the comparability of diabetes mortality statistics in the U.S. and Mexico. Diabetes Care 2008;31(3):451-8.
109. Najafi F, Dobson AJ, Jamrozik K. Is mortality from heart failure increasing in Australia? An analysis of official data on mortality for 1997-2003. Bull World Health Organ 2006;84(9):722-8.
110. Nam CB, Hummer RA, Rogers RG. Underlying and multiple causes of death related to smoking. Population Research and Policy Review 1994;13(3):305-325.
111. Nashold RD, Naor EM. Alcohol-related deaths in Wisconsin: the impact of alcohol on mortality. Am J Public Health 1981;71(11):1237-41.
112. Nath U, Thomson R, Wood R, et al. Population based mortality and quality of death certification in progressive supranuclear palsy (Steele-Richardson-Olszewski syndrome). J Neurol Neurosurg Psychiatry 2005;76(4):498-502.
113. Nizamo H, Meyrowitsch DW, Zacarias E, Konradsen F. Mortality due to injuries in Maputo City, Mozambique. Int J Inj Contr Saf Promot 2006;13(1):1-6.
114. Nizard A, Munozperez F. Alcohol, tobacco, and mortality in France since 1950 - An estimate of the annual numbers of deaths. Population 1993;48(3):571-607.
115. Nwancha AB, Alvarado E, Ma J, Gillum RF, Hughes K. Atherosclerotic Peripheral Artery Disease in the United States: Gender and ethnic variation in a multiple cause-of-death analysis. Vasc Endovascular Surg 2020;54(6):482-486.
116. Obi J, Mehari A, Gillum R. Mortality related to chronic obstructive pulmonary disease and co-morbidities in the United States, A multiple causes of death analysis. COPD 2018;15(2):200-205.
117. Ochi JW, Melton LJ, 3rd, Palumbo PJ, Chu CP. A population-based study of diabetes mortality. Diabetes Care 1985;8(3):224-9.
118. Okuda T, Wang Z, Lapan S, Fowler DR. Bathtub drowning: An 11-year retrospective study in the state of Maryland. Forensic Sci Int 2015;253:64-70.
119. Olson AL, Swigris JJ, Sprunger DB, et al. Rheumatoid arthritis-interstitial lung disease-associated mortality. Am J Respir Crit Care Med 2011;183(3):372-8.
120. Owe JF, Daltveit AK, Gilhus NE. Does myasthenia gravis provide protection against cancer? Acta Neurol Scand 2006;113:33-36.
121. Pabich SB, N. Trends in hip fracture mortality in Wisconsin and the United States, 1999-2017. WMJ 2020.
122. Pacheco AG, Tuboi SH, Faulhaber JC, Harrison LH, Schechter M. Increase in non-AIDS related conditions as causes of death among HIV-infected individuals in the HAART era in Brazil. PLoS One 2008;3(1):e1531.
123. Paik JM, Henry L, De Avila L, et al. Mortality related to nonalcoholic fatty liver disease is increasing in the United States. Hepatol Commun 2019;3(11):1459-1471.
124. Paik JM, Golabi P, Biswas R, et al. Nonalcoholic fatty liver disease and alcoholic liver disease are major drivers of liver mortality in the United States. Hepatol Commun 2020;4(6):890-903.
125. Palladino C, Climent FJ, De Jose MI, et al. Causes of death in pediatric patients vertically infected by the Human Immunodeficiency Virus Type 1 in Madrid, Spain, rrom 1982 to mid-2009. Pediatric Infectious Disease Journal 2011;30(6):495-500.
126. Park CB, Yokoyama E, Tokuyama GH. Medical conditions at death among the Caucasian and Japanese elderly in Hawaii - analysis of multiple causes of death, 1976-78. Journal of Clinical Epidemiology 1991;44(6):519-530.
127. Parks SE, Kegler SR, Annest JL, Mercy JA. Characteristics of fatal abusive head trauma among children in the USA: 2003-2007: an application of the CDC operational case definition to national vital statistics data. Inj Prev 2012;18(3):193-9.
128. Paulozzi LJ, Budnitz DS, Xi Y. Increasing deaths from opioid analgesics in the United States. Pharmacoepidemiol Drug Saf 2006;15(9):618-27.
129. Pechholdová M. Multiple cause-of-death data in the Czech Republic: An exploratory analysis. Demografie 2014;56(4):335-346.
130. Pechholdova M. Sepsis-related mortality in the Czech Republic: multiple causes of death analysis. Epidemiologie, Mikrobiologie, Imunologie. 66(2):73-79, Summer 2017. 2017.
131. Peng JK, Higginson IJ, Gao W. Place of death and factors associated with hospital death in patients who have died from liver disease in England: a national population-based study. Lancet Gastroenterol Hepatol 2019;4(1):52-62.
132. Pillutla P, Sherry KD, Foster E. Mortality associated with adult congenital heart disease: Trends in the US population from 1979 to 2005. Am Heart J 2009;158(5):874-879.
133. Pinheiro FA, Souza DC, Sato EI. A Study of Multiple Causes of Death in Rheumatoid Arthritis. J Rheumatol 2015;42(12):2221-8.
134. Polednak AP. Trend (1999-2009) in U.S. death rates from myelodysplastic syndromes: utility of multiple causes of death in surveillance. Cancer Epidemiol 2013;37(5):569-74.
135. Polednak AP. Trend in rates for deaths with mention of schizophrenia on death certificates of US residents, 1999-2010. Soc Psychiatry Psychiatr Epidemiol 2014;49(7):1083-91.
136. Polednak AP. Surveillance of US death rates from chronic diseases related to excessive alcohol use. Alcohol and Alcoholism 2015;51(1):54-62.
137. Poston Jr DL, Min H. The multinomial regression modeling of the cause-of-death mortality of the oldest old in the U.S. Journal of Modern Applied Statistical Methods 2008;7(2):597-606.
138. Prescott E, Lange P, Vestbo J. Chronic mucus hypersecretion in COPD and death from pulmonary infection. Eur Respir J 1995;8(8):1333-8.
139. Rasmussen SA, Yang Q, Friedman JM. Mortality in neurofibromatosis 1: an analysis using U.S. death certificates. Am J Hum Genet 2001;68(5):1110-8.
140. Riihimaki M, Thomsen H, Brandt A, Sundquist J, Hemminki K. Death causes in breast cancer patients. Ann Oncol 2012;23(3):604-610.
141. Roaeid RB, Kablan AA. Diabetes mortality and causes of death in Benghazi: a 5-year retrospective analysis of death certificates. East Mediterr Health J 2010;16(1):65-9.
142. Rocha MS, de Oliveira GP, Aguiar FP, Saraceni V, Pinheiro RS. What are the causes of death of patients with tuberculosis: multiple causes of death in a cohort of cases and a research proposal of presumed causes. Cad Saude Publica 2015;31(4):709-721.
143. Rodriguez F, Blum MR, Falasinnu T, et al. Diabetes-attributable mortality in the United States from 2003 to 2016 using a multiple-cause-of-death approach. Diabetes Res Clin Pract 2019;148:169-178.
144. Roxburgh A, Lappin J. MDMA-related deaths in Australia 2000 to 2018. Int J Drug Policy 2020;76:102630.
145. Rudd KE, Johnson SC, Agesa KM, et al. Global, regional, and national sepsis incidence and mortality, 1990&#x2013;2017: analysis for the Global Burden of Disease Study. The Lancet 2020;395(10219):200-211.
146. Ruhm CJ. Drug involvement in fatal overdoses. SSM Popul Health 2017;3:219-226.
147. Ruhm CJ. Corrected US opioid-involved drug poisoning deaths and mortality rates, 1999-2015. Addiction 2018;113(7):1339-1344.
148. Rushton L. Use of multiple causes of death in the analysis of occupational cohorts--an example from the oil industry. Occup Environ Med 1994;51(11):722-9.
149. Ruzicka LT, Choi CY, Sadkowsky K. Medical disorders of suicides in Australia: analysis using a multiple-cause-of-death approach. Soc Sci Med 2005;61(2):333-41.
150. Salive ME, Satterfield S, Ostfeld AM, Wallace RB, Havlik RJ. Disability and cognitive impairment are risk factors for pneumonia-related mortality in older adults. Public Health Rep 1993;108(3):314-22.
151. Sampaio VS, Rodrigues MGA, Silva L, et al. Social, demographic, health care and co-morbidity predictors of tuberculosis mortality in Amazonas, Brazil: a multiple cause of death approach. PLoS One 2020;15(1):e0218359.
152. Santo AH, Pinheiro CE, Jordani MS. [Aids as underlying and associated causes of death, State of S. Paulo, Brazil, 1998]. Rev Saude Publica 2000;34(6):581-8.
153. Santo AH, Pinheiro CE, Jordani MS. [Multiple-causes-of-death related to tuberculosis in the State of São Paulo, Brazil, 1998]. Rev Saude Publica 2003;37(6):714-21.
154. Santo AH. Deaths attributed to multiple causes and involving tuberculosis in the state of Rio de Janeiro Brazil between 1999 and 2001. J Bras Pneumol 2006;32(6):544-52.
155. Santo AH, Souza JM, Pinheiro CE, Souza DC, Sato EI. Trends in dermatomyositis- and polymyositis-related mortality in the state of São Paulo, Brazil, 1985-2007: multiple cause-of-death analysis. BMC Public Health 2010;10:597.
156. Santo AH. Causes of death and mortality trends related to hemophilia in Brazil, 1999 to 2016. Hematol Transfus Cell Ther 2020.
157. Seuc AH, Fernandez L, Mirabal M, Rodriguez A, Rodriguez CA. Cuban Application of Two Methods for Analyzing Multiple Causes of Death. MEDICC Rev 2018;20(3):30-35.
158. Sleeman KE, Ho YK, Verne J, et al. Place of death, and its relation with underlying cause of death, in Parkinson's disease, motor neurone disease, and multiple sclerosis: a population-based study. Palliat Med 2013;27(9):840-6.
159. Sohn AH, Lumbiganon P, Kurniati N, et al. Determining standardized causes of death of infants, children, and adolescents living with HIV in Asia. AIDS 2020;34(10):1527-1537.
160. Souza DC, Santo AH, Sato EI. Mortality profile related to systemic lupus erythematosus: a multiple cause-of-death analysis. J Rheumatol 2012;39(3):496-503.
161. Speizer FE, Trey C, Parker P. The uses of multiple causes of death data to clarify changing patterns of cirrhosis mortality in Massachusetts. Am J Public Health 1977;67(4):333-6.
162. Stallard E. Underlying and Multiple Case Mortality Advanced Ages: United States 1980-1998. North American Actuarial Journal 2002;6(3):64-87.
163. Stares J, Kosatsky T. Hypothermia as a cause of death in British Columbia, 1998-2012: a descriptive assessment. CMAJ Open 2015;3(4):E352-8.
164. Sterling TDR, W. L.; Weinkam, J. J. Bias in the attribution of lung cancer as cause of death and its possible consequences for calculating smoking-related risks. Epidemiology 1992.
165. Suligoi B, Virdone S, Taborelli M, et al. Excess mortality related to circulatory system diseases and diabetes mellitus among Italian AIDS patients vs. non-AIDS population: a population-based cohort study using the multiple causes-of-death approach. BMC Infectious Diseases 2018;18.
166. Sweitzer K, Stallones L. Significant contributing causes of cancer deaths among Hispanics in Colorado, USA, 1983-1992. Cad Saude Publica 1998;14 Suppl 3:187-91.
167. Swigris JJ, Olson AL, Huie TJ, et al. Sarcoidosis-related mortality in the United States from 1988 to 2007. Am J Respir Crit Care Med 2011;183(11):1524-30.
168. Takamori A, Takahashi I, Kasagi F, et al. Mortality analysis of the Life Span Study (LSS) cohort taking into account multiple causes of death indicated in death certificates. Radiat Res 2017;187(1):20-31.
169. Tamashiro H, Akagi H, Arakaki M, Futatsuka M, Roht LH. Causes of death in Minamata disease: analysis of death certificates. Int Arch Occup Environ Health 1984;54(2):135-46.
170. Thomas G, Mancini J, Jourde-Chiche N, et al. Mortality associated with systemic lupus erythematosus in France assessed by multiple-cause-of-death analysis. Arthritis Rheumatol 2014;66(9):2503-11.
171. Turner C, Chandrakumar D, Rowe C, et al. Cross-sectional cause of death comparisons for stimulant and opioid mortality in San Francisco, 2005-2015. Drug Alcohol Depend 2018;185:305-312.
172. Van Natta P, Malin H, Bertolucci D, Kaelber C. The influence of alcohol abuse as a hidden contributor to mortality. Alcohol 1985;2(3):535-9.
173. Villela PB, Klein CH, Oliveira GMM. Cerebrovascular and hypertensive diseases as multiple causes of death in Brazil from 2004 to 2013. Public Health 2018;161:36-42.
174. Vinnard C, Longworth S, Mezochow A, et al. Deaths related to Nontuberculous mycobacterial infections in the United States, 1999-2014. Ann Am Thorac Soc 2016;13(11):1951-1955.
175. Wattigney WA, Mensah GA, Croft JB. Increased atrial fibrillation mortality: United States, 1980-1998. Am J Epidemiol 2002;155(9):819-26.
176. White MC. Mortality associated with nosocomial infections: analysis of multiple cause-of-death data. J Clin Epidemiol 1993;46(1):95-100.
177. White MC, Portillo CJ. Tuberculosis mortality associated with AIDS and drug or alcohol abuse: analysis of multiple cause-of-death data. Public Health 1996;110(3):185-9.
178. White AM, Castle IP, Hingson RW, Powell PA. Using death certificates to explore changes in alcohol-related mortality in the United States, 1999 to 2017. Alcohol Clin Exp Res 2020;44(1):178-187.
179. Whiteside YO, Selik R, An Q, et al. Comparison of rates of death having any death-certificate mention of heart, kidney, or liver disease among persons diagnosed with HIV Infection with those in the General US Population, 2009-2011. Open AIDS J 2015;9:14-22.
180. Wickramasekaran RN, Sorvillo F, Kuo T. Legionnaires' disease and associated comorbid conditions as causes of death in the U.S., 2000-2010. Public Health Rep 2015;130(3):222-9.
181. Wilkins K, Parsons GF, Gentleman JF, Forbes WF. Deaths due to dementia: An analysis of multiple-cause-of-death data. Chronic Dis Can 1999;20(1):26-35.
182. Wong O, Rockette HE, Redmond CK, Heid M. Evaluation of multiple causes of death in occupational mortality studies. Journal of Chronic Diseases 1978;31(3):183-193.
183. Wrigley JM, Nam CB. Underlying versus multiple causes of death - Effects on interpreting cancer mortality differentials by age, sex, and race. Population Research and Policy Review 1987;6(2):149-160.
184. Yang Q, McDonnell SM, Khoury MJ, Cono J, Parrish RG. Hemochromatosis-associated mortality in the United States from 1979 to 1992: an analysis of Multiple-Cause Mortality Data. Ann Intern Med 1998;129(11):946-53.
185. Yashin AI, Ukraintseva SV, Akushevich IV, et al. Trade-off between cancer and aging: what role do other diseases play? Evidence from experimental and human population studies. Mech Ageing Dev 2009;130(1-2):98-104.
186. Yen EY, Singh RR. Lupus-An Unrecognized Leading Cause of Death in Young Females: A Population-Based Study Using Nationwide Death Certificates, 2000-2015. Arthritis & Rheumatology 2018;70(8):1251-1255.
187. Yoon YH, Stinson FS, Yi HY, Dufour MC. Accidental alcohol poisoning mortality in the United States, 1996-1998. Alcohol Res Health 2003;27(1):110-8.
188. Yoon YH, Chen CM, Yi HY, Moss HB. Effect of comorbid alcohol and drug use disorders on premature death among unipolar and bipolar disorder decedents in the United States, 1999 to 2006. Compr Psychiatry 2011;52(5):453-64.
189. Zargar AH, Wani AI, Masoodi SR, et al. Causes of mortality in diabetes mellitus: data from a tertiary teaching hospital in India. Postgrad Med J 2009;85(1003):227-32.
190. Ziade N, Jougla E, Coste J. Population-level impact of osteoporotic fractures on mortality and trends over time: a nationwide analysis of vital statistics for France, 1968-2004. Am J Epidemiol 2010;172(8):942-51.
191. Zoppini G, Fedeli U, Gennaro N, et al. Mortality from chronic liver diseases in diabetes. Am J Gastroenterol 2014;109(7):1020-5.
192. Adair T, Lopez AD. The role of overweight and obesity in adverse cardiovascular disease mortality trends: an analysis of multiple cause of death data from Australia and the USA. BMC Med 2020;18(1):199.
193. Bah S. Multiple causes-of-death statistics in South Africa: their changing profile over the period 1997 to 2001 and utility in the era of HIV/AIDS. The South African Journal of Epidemiology and Infection 2005.
194. Baker SP, Brady JE, Shanahan DF, Li G. Aviation-related injury morbidity and mortality: data from U.S. health information systems. Aviat Space Environ Med 2009;80(12):1001-5.
195. Balkau B, Papoz L. Certification of cause of death in French diabetic patients. J Epidemiol Community Health 1992;46(1):63-5.
196. Barbieri M, Désesquelles A, Egidi V, et al. Obesity-related mortality in France, Italy, and the United States: a comparison using multiple cause-of-death analysis. Int J Public Health 2017;62(6):623-629.
197. Barreto SM, Passos VM, Almeida SK, Assis TD. The increase of diabetes mortality burden among Brazilian adults. Rev Panam Salud Publica 2007;22(4):239-45.
198. Burke JF, Lisabeth LD, Brown DL, Reeves MJ, Morgenstern LB. Determining stroke's rank as a cause of death using multicause mortality data. Stroke 2012;43(8):2207-11.
199. Buschner A, Grunwald-Muhlberger A. Influence of methodological changes on unicausal cause-of-death statistics and potentials of a multicausal data basis. Bundesgesundheitsblatt-Gesundheitsforschung-Gesundheitsschutz 2019;62(12):1476-1484.
200. Bustamante-Montes LP, Alvarez-Solorza I, Valencia AD, et al. Applicability of the analysis by multiple cause of death by cervical cancer: The experience in Mexico. Cien Saude Colet 2011;16(12):4815-4821.
201. Cascão AM, Costa AJ, Kale PL. [Quality of mortality information in a diabetes cohort--State of Rio de Janeiro, 2000 to 2003]. Rev Bras Epidemiol 2012;15(1):134-42.
202. Castle IJ, Yi HY, Hingson RW, White AM. State variation in underreporting of alcohol involvement on death certificates: motor vehicle traffic crash fatalities as an example. J Stud Alcohol Drugs 2014;75(2):299-312.
203. Coeli CM, Ferreira LG, Drbal Md Mde M, et al. [Diabetes mellitus mortality among elderly as an underlying or secondary cause of death]. Rev Saude Publica 2002;36(2):135-40.
204. Cohen J, Steinitz R. Underlying and contributory causes of death of adult males in two districts. J Chronic Dis 1969;22(1):17-24.
205. Crews DE. Multiple causes of death and the epidemiological transition in American Samoa. Soc Biol 1988;35(3-4):198-213.
206. Cutter GR, Zimmerman J, Salter AR, et al. Causes of death among persons with multiple sclerosis. Mult Scler Relat Disord 2015;4(5):484-490.
207. da Silva EM, e Silva GA. Asthma-related mortality in the city of Rio de Janeiro, Brazil, 2000-2009: A multicausal analysis. Cad Saude Publica 2013;29(4):667-680.
208. de Oliveira BZ, Gotlieb SLD, Laurenti R, Jorge MHPM. Mortality due to hypertension in women: A multiple cause analysis. Revista Brasileira de Epidemiologia 2009;12(4):556-565.
209. Dembling B. Mental disorder as a contributing cause of death in the U.S. in 1992. Psychiatr Serv 1997;48(1):45.
210. Désesquelles AD, E.; Mésle, F.; Salvatore, M A. Cause-specific mortality analysis: Is the underlying cause of death sufficient? Revue Quetelet 2014.
211. Désesquelles A, Gamboni A, Demuru E, et al. We only die once... but from how many causes? Population and Societies 2016;534(1):1-4.
212. Doi Y, Yokoyama T, Nakamura Y, et al. How can the national burden of Parkinson's disease comorbidity and mortality be estimated for the Japanese population? J Epidemiol 2011;21(3):211-6.
213. Domingues CSB, Waldman EA. Causes of death among people living with AIDS in the pre- and post-HAART eras in the city of S(a)over-tildeo Paulo, Brazil. PLoS One 2014;9(12).
214. Doney BC, Miller WE, Hale JM, Syamlal G. Estimation of the number of workers exposed to respirable crystalline silica by industry: Analysis of OSHA compliance data (1979-2015). Am J Ind Med 2020;63(6):465-477.
215. Dorn H MI. Uses and significance of multiple cause tabulations for mortality statistics. AJPH 1964;54(3).
216. D'Souza MJ, Li RC, Wentzien DE. Delaware's 1999-2017 leading causes of death information illustrates its obesity and obesity-related life-limiting disease burdens. Res Health Sci 2019;4(4):327-346.
217. Duncan ME, Goldacre MJ. Mortality trends for tuberculosis and sarcoidosis in English populations, 1979-2008. Int J Tuberc Lung Dis 2012;16(1):38-42.
218. Duncan ME, Goldacre MJ. Certification of deaths from diabetes mellitus and obesity in England: trends into the twenty-first century. J Public Health (Oxf) 2013;35(2):293-7.
219. Egidi V, Salvatore MA, Rivellini G, D'Angelo S. A network approach to studying cause-of-death interrelations. Demogr Res 2018;38(1):373-400.
220. Esposito DH, Holman RC, Haberling DL, et al. Baseline estimates of diarrhea-associated mortality among United States children before rotavirus vaccine introduction. Pediatr Infect Dis J 2011;30(11):942-7.
221. Fedeli U, Schievano E, Lisiero M, et al. Descriptive epidemiology of chronic liver disease in northeastern Italy: an analysis of multiple causes of death. Popul Health Metr 2013;11(1):20.
222. Ferreira AF, de Souza EA, Lima MDS, et al. [Mortality from leprosy in highly endemic contexts: integrated temporal-spatial analysis in BrazilMortalidad por lepra en zonas de alta endemicidad: análisis espacio-temporal integrado en Brasil]. Rev Panam Salud Publica 2019;43:e87.
223. Fife D. Injuries and deaths among elderly persons. Am J Epidemiol 1987;126(5):936-41.
224. Fife D, Rappaport E. What role do injuries play in the deaths of old people? Accid Anal Prev 1987;19(3):225-30.
225. Fink AK, German RR, Heron M, et al. Impact of using multiple causes of death codes to compute site-specific, death certificate-based cancer mortality statistics in the United States. Cancer Epidemiol 2012;36(1):22-8.
226. Flaten TP. Mortality from dementia in Norway, 1969-83. J Epidemiol Community Health 1989;43(3):285-9.
227. Ford MM, Desai PS, Maduro G, Laraque F. Neighborhood inequalities in Hepatitis C mortality: Spatial and temporal patterns and associated factors. J Urban Health 2017;94(5):746-755.
228. Fuhrman C, Jougla E, Nicolau J, Eilstein D, Delmas MC. Deaths from chronic obstructive pulmonary disease in France, 1979-2002: A multiple cause analysis. Thorax 2006;61(11):930-934.
229. Furukawa TS, Santo AH, Mathias TA. Multiple causes of death related to cerebrovascular diseases in the State of Parana, Brazil. Rev Bras Epidemiol 2011;14(2):231-9.
230. Garshick E, Kelley A, Cohen SA, et al. A prospective assessment of mortality in chronic spinal cord injury. Spinal Cord 2005;43(7):408-16.
231. Gaui EN, Klein CH, De Oliveira GMM. Mortality due to heart failure: Extended analysis and temporal trend in three states of Brazil. Arq Bras Cardiol 2010;94(1):52-58.
232. Gillum RF, Obisesan TO. Differences in mortality associated with dementia in U.S. blacks and whites. J Am Geriatr Soc 2011;59(10):1823-8.
233. Glei DA, Preston SH. Estimating the impact of drug use on US mortality, 1999-2016. PLoS One 2020;15(1):e0226732.
234. Goldacre MJ. Cause-specific mortality: understanding uncertain tips of the disease iceberg. J Epidemiol Community Health 1993;47(6):491-6.
235. Goldacre MJ, Duncan ME, Cook-Mozaffari P, Griffith M. Trends in mortality rates comparing underlying-cause and multiple-cause coding in an English population 1979-1998. J Public Health Med 2003;25(3):249-53.
236. Goldacre MJ, Duncan M, Cook-Mozaffari P, Griffith M. Mortality rates for common respiratory diseases in an English population 1979-1998: artefact and substantive trends. J Public Health (Oxf) 2004;26(1):8-12.
237. Goldacre MJ, Duncan M, Cook-Mozaffari P, Griffith M. Trends in mortality for cancers, comparing multiple- and underlying-cause rates, in an English population 1979-1999. Br J Cancer 2004;90(5):1019-1021.
238. Goldacre MJ, Duncan ME, Cook-Mozaffari P, Neil HA. Trends in mortality rates for death-certificate-coded diabetes mellitus in an English population 1979-99. Diabet Med 2004;21(8):936-9.
239. Goldacre MJ, Duncan M, Griffith M, Cook-Mozaffari P. Alcohol as a certified cause of death in a 'middle England' population 1979-1999: database study. J Public Health (Oxf) 2004;26(4):343-6.
240. Goldacre MJ, Duncan ME, Griffith M, Cook-Mozaffari P. Psychiatric disorders certified on death certificates in an English population. Soc Psychiatry Psychiatr Epidemiol 2006;41(5):409-14.
241. Goldacre MJ, Duncan M, Cook-Mozaffari P, Griffith M, Travis S. Inflammatory bowel disease, peptic ulcer and diverticular disease as certified causes of death in an English population 1979-2003. Eur J Gastroenterol Hepatol 2008;20(2):96-103.
242. Goldacre MJ, Duncan M, Griffith M, Rothwell PM. Mortality rates for stroke in England from 1979 to 2004: trends, diagnostic precision, and artifacts. Stroke 2008;39(8):2197-203.
243. Goldacre MJ, Duncan M, Griffith M, Turner MR. Trends in death certification for multiple sclerosis, motor neuron disease, Parkinson's disease and epilepsy in English populations 1979-2006. J Neurol 2010;257(5):706-15.
244. Goldacre MJ, Duncan ME, Griffith M. Death rates for asthma in English populations 1979-2007: comparison of underlying cause and all certified causes. Public Health 2012;126(5):386-93.
245. Goldstein E, Lipsitch M. The relation between prescribing of different antibiotics and rates of mortality with sepsis in US adults. BMC Infect Dis 2020;20(1):169.
246. Gonzalez LF, Jo AHS, Garcia CAR. Weighted mortality method according to multiple causes of death. Finlay 2019;9(3):197-209.
247. Greenlund SF, Croft JB, Kobau R. Epilepsy by the numbers: Epilepsy deaths by age, race/ethnicity, and gender in the United States significantly increased from 2005 to 2014. Epilepsy Behav 2017;69:28-30.
248. Grippo F, Navarra S, Orsi C, et al. The role of COVID-19 in the death of SARS-CoV-2-Positive patients: A study vased on death certificates. J Clin Med 2020;9(11).
249. Hansell AL, Walk JA, Soriano JB. What do chronic obstructive pulmonary disease patients die from? A multiple cause coding analysis. Eur Respir J 2003;22(5):809-14.
250. Harding JL, Shaw JE, Peeters A, et al. Mortality trends among people with type 1 and type 2 diabetes in Australia: 1997-2010. Diabetes Care 2014;37(9):2579-86.
251. Harzke AJ, Baillargeon JG, Kelley MF, et al. HCV-related mortality among male prison inmates in Texas, 1994-2003. Ann Epidemiol 2009;19(8):582-9.
252. Huang JY, Bristow B, Shafir S, Sorvillo F. Coccidioidomycosis-associated Deaths, United States, 1990-2008. Emerg Infect Dis 2012;18(11):1723-8.
253. Hummer RA, Nam CB, Rogers RG. Adult mortality differentials associated with cigarette smoking in the USA. Population Research and Policy Review 1998;17(3):285-304.
254. Jansson B, Johansson LA, Rosen M, Svanstrom L. National adaptations of the ICD rules for classification--a problem in the evaluation of cause-of-death trends. Journal of Clinical Epidemiology. 50(4):367-75, 1997 Apr. 1997.
255. Jansson B, Ahmed N. Epilepsy and injury mortality in Sweden--the importance of changes in coding practice. Seizure 2002;11(6):361-70.
256. Jeganathan N, Smith RA, Sathananthan M. Mortality Trends of Idiopathic Pulmonary Fibrosis in the United States from 2004 through 2017. Chest 2020;159(1):228-238.
257. Jowching E. Multiple cause-of-death analysis of hypertension-related mortality in New-York-State. Public Health Reports 1987;102(3):329-335.
258. Karkkainen M, Nurmi H, Kettunen HP, et al. Underlying and immediate causes of death in patients with idiopathic pulmonary fibrosis. BMC Pulm Med 2018;18(1):69.
259. Kaufmann RB, Staes CJ, Matte TD. Deaths related to lead poisoning in the United States, 1979-1998. Environ Res 2003;91(2):78-84.
260. Khou V, De La Mata NL, Morton RL, Kelly PJ, Webster AC. Cause of death for people with end-stage kidney disease withdrawing from treatment in Australia and New Zealand. Nephrol Dial Transplant 2020;36(8):1527-1537.
261. Kodadhala V, Obi J, Wessly P, Mehari A, Gillum RF. Asthma-related mortality in the United States, 1999 to 2015: A multiple causes of death analysis. Ann Allergy Asthma Immunol 2018;120(6):614-619.
262. Kosan Z, Bedir B, Yilmaz S, et al. An evaluation of the infant mortality rate in 2014 and 2015 in Northeastern Anatolia. Konuralp Tip Dergisi 2019;11(1):76-81.
263. Krueger DE. Hypertensive and chronic respiratory disease mortality - confirmation of trends by multiple cause of death data. Public Health Reports 1966;81(2):197-+.
264. Kuller L, Seltser R, Paffenbarger RS, Jr., Krueger DE. Trends in cerebrovascular disease mortality based on multiple cause tabulation of death certificates 1930-1960. A comparison of trends in Memphis and Baltimore. Am J Epidemiol 1968;88(3):307-17.
265. Lanska DJ, Peterson PM. Geographic variation in reporting of stroke deaths to underlying or contributing causes in the United States. Stroke 1995;26(11):1999-2003.
266. Laribi S, Aouba A, Resche-Rigon M, et al. Trends in death attributed to myocardial infarction, heart failure and pulmonary embolism in Europe and Canada over the last decade. Qjm 2014;107(10):813-20.
267. Larsen JP, Kvale G, Aarli JA. Multiple sclerosis and mortality statistics. Acta Neurol Scand 1985;71(3):237-241.
268. Larson TC, Kaye W, Mehta P, Horton DK. Amyotrophic Lateral Sclerosis mortality in the United States, 2011-2014. Neuroepidemiology 2018;51(1-2):96-103.
269. Laurenti R. Analysis of mortality, based on basic cause of death and on multiple causes. Rev Saude Publica 1974;8(4):421-435.
270. Leung J, Bialek SR, Marin M. Trends in varicella mortality in the United States: Data from vital statistics and the national surveillance system. Hum Vaccin Immunother 2015;11(3):662-8.
271. Lin YP, Lu TH. Trends in death rate from diabetes according to multiple-cause-of-death differed from that according to underlying-cause-of-death in Taiwan but not in the United States, 1987-2007. J Clin Epidemiol 2012;65(5):572-6.
272. Lin JJ, Liang FW, Li CY, Lu TH. Leading causes of death among decedents with mention of schizophrenia on the death certificates in the United States. Schizophr Res 2018;197:116-123.
273. Lindahl BI, Johansson LA. Multiple cause-of-death data as a tool for detecting artificial trends in the underlying cause statistics: a methodological study. Scand J Soc Med 1994;22(2):145-58.
274. Lindeus M, Englund M, Kiadaliri AA. Educational inequalities in fracture-related mortality using multiple cause of death data in the Skane region, Sweden. Scand J Public Health 2020;48(1):72-79.
275. Lu TH, Walker S, Johansson LA, Huang CN. An international comparison study indicated physicians' habits in reporting diabetes in part I of death certificate affected reported national diabetes mortality. J Clin Epidemiol 2005;58(11):1150-7.
276. Lu TH, Hsu PY, Bjorkenstam C, Anderson RN. Certifying diabetes-related cause-of-death: a comparison of inappropriate certification statements in Sweden, Taiwan and the USA. Diabetologia 2006;49(12):2878-81.
277. Lu TH, Lin JJ. Using multiple-cause-of-death data as a complement of underlying-cause-of-death data in examining mortality differences in psychiatric disorders between countries. Soc Psychiatry Psychiatr Epidemiol 2010;45(8):837-42.
278. Lu TH, Hsiao A, Chang PC, et al. Counting injury deaths: a comparison of two definitions and two countries. Inj Prev 2013;21(e1):e127-32.
279. Madsen M, Davidsen M, Rasmussen S, Abildstrom SZ, Osler M. The validity of the diagnosis of acute myocardial infarction in routine statistics: a comparison of mortality and hospital discharge data with the Danish MONICA registry. J Clin Epidemiol 2003;56(2):124-30.
280. Makela P. Alcohol-related mortality by age and sex and its impact on life expectancy - Estimates based on the Finnish death register. Eur J Public Health 1998;8(1):43-51.
281. Manderbacka K, Arffman M, Lyytikainen O, Sajantila A, Keskimaki I. What really happened with pneumonia mortality in Finland in 2000-2008?: a cohort study. Epidemiol Infect 2013;141(4):800-4.
282. Mannino DM, Brown C, Giovino GA. Obstructive lung disease deaths in the United States from 1979 through 1993. An analysis using multiple-cause mortality data. Am J Respir Crit Care Med 1997;156(3 Pt 1):814-8.
283. Manton KG. Sex and race specific mortality differentials in multiple cause of death data. Gerontologist 1980.
284. Manton KG, Patrick, C. H., Stallard, E. Population Impact of Mortality Reduction: The Effects of Elimination of Major Causes of Death on the 'Saved' Population. International Journal of Epidemiology 1980.
285. Manton KGS, E. Mortality of the chronically impaired. Demography 1980.
286. Manton KG, Stallard E. Temporal trends in U. S. multiple cause of death mortality data: 1968 to 1977. Demography 1982;19(4):527-47.
287. Manton KGB, H. M. CVD mortality, 1968-1978: observations and implications. Stroke 1984.
288. Manton KG. Cause specific mortality patterns among the oldest old: multiple cause of death trends 1968 to 1980. J Gerontol 1986;41(2):282-9.
289. Manton KG, Myers GC. Recent trends in multiple-caused mortality 1968 to 1982: Age and cohort components. Population Research and Policy Review 1987;6(2):161-176.
290. Manton KG, Wrigley JM, Cohen HJ, Woodbury MA. Cancer mortality, aging, and patterns of comorbidity in the United States: 1968 to 1986. J Gerontol 1991;46(4):S225-34.
291. Marin M, Zhang JX, Seward JF. Near elimination of varicella deaths in the US after implementation of the vaccination program. Pediatrics 2011;128(2):214-20.
292. Markush RE, Siegel, D. C. Prevalence at death. A new method for deriving rates for specific diseases. Am J Public Health 1968.
293. Martins EF, Almeida PF, Paixão CO, Bicalho PG, Errico LS. [Multiple causes of maternal mortality related to abortion in Minas Gerais State, Brazil, 2000-2011]. Cad Saude Publica 2017;33(1):e00133115.
294. Martins-Melo FR, Alencar CH, Ramos AN, Jr., Heukelbach J. Epidemiology of mortality related to Chagas' disease in Brazil, 1999-2007. PLoS Negl Trop Dis 2012;6(2):e1508.
295. Martins-Melo FR, Ramos AN, Alencar CH, Heukelbach J. Mortality related to Chagas disease and HIV/AIDS coinfection in Brazil. Journal of Tropical Medicine 2012.
296. Martins-Melo FR, da Silveira Lima M, Alencar CH, Ramos Jr AN, Heukelbach J. Epidemiological patterns of mortality due to visceral leishmaniasis and HIV/AIDS co-infection in Brazil, 2000-2011. Transactions of the Royal Society of Tropical Medicine and Hygiene 2014;108(6):338-347.
297. Martins-Melo FR, Pinheiro MC, Ramos AN, Jr., et al. Trends in schistosomiasis-related mortality in Brazil, 2000-2011. Int J Parasitol 2014;44(14):1055-62.
298. Martins-Melo FR, Ramos AN, Alencar CH, Lima MS, Heukelbach J. Epidemiology of soil-transmitted helminthiases-related mortality in Brazil. Parasitology 2017;144(5):669-679.
299. Maudsley G, Hutton JL, Pharoah PO. Cause of death in cerebral palsy: a descriptive study. Arch Dis Child 1999;81(5):390-4.
300. McCoy L, Sorvillo F, Simon P. Varicella-related mortality in California, 1988-2000. Pediatr Infect Dis J 2004;23(6):498-503.
301. McCoy L, Redelings M, Sorvillo F, Simon P. A multiple cause-of-death analysis of asthma mortality in the United States, 1990-2001. J Asthma 2005;42(9):757-63.
302. Meyers DJ, Hood ME, Stopka TJ. HIV and hepatitis C mortality in Massachusetts, 2002-2011: spatial cluster and trend analysis of HIV and HCV using multiple cause of death. PLoS One 2014;9(12):e114822.
303. Midgard R, Albrektsen G, Riise T, Kvale G, Nyland H. Prognostic factors for survival in multiple sclerosis: a longitudinal, population based study in More and Romsdal, Norway. Journal of Neurology, Neurosurgery & Psychiatry. 58(4):417-21, 1995 Apr. 1995.
304. Milham S, Jr. Using multiple cause of death coding in occupational mortality studies. Am J Ind Med 1988;14(3):341-4.
305. Mitratza M, Kunst AE, Harteloh PPM, Nielen MMJ, Klijs B. Prevalence of diabetes mellitus at the end of life: An investigation using individually linked cause-of-death and medical register data. Diabetes Res Clin Pract 2020;160:108003.
306. Moreno-Betancur M, Sadaoui H, Piffaretti C, Rey G. Survival Analysis with multiple causes of death extending the Competing Risks Model. Epidemiology 2017;28(1):12-19.
307. Moussa MA, El Sayed AM, Sugathan TN, Khogali MM, Verma D. Analysis of underlying and multiple-cause mortality data. Genus 1992;48(1-2):89-105.
308. Murdoch DR, Love MP, Robb SD, et al. Importance of heart failure as a cause of death. Changing contribution to overall mortality and coronary heart disease mortality in Scotland 1979-1992. Eur Heart J 1998;19(12):1829-35.
309. Nembhard WN, Pathak EB, Schocken DD. Racial/ethnic disparities in mortality related to congenital heart defects among children and adults in the United States. Ethn Dis 2008;18(4):442-9.
310. Newman JM, DeStefano F, Valway SE, German RR, Muneta B. Diabetes-associated mortality in Native Americans. Diabetes Care 1993;16(1):297-9.
311. Ning P, Schwebel DC, Chu H, Zhu M, Hu G. Changes in reporting for unintentional injury deaths, United States of America. Bull World Health Organ 2019;97(3):190-199.
312. Nordstrom DL, Yokoi-Shelton ML, Zosel A. Using multiple cause-of-death data to improve surveillance of drug-related mortality. J Public Health Manag Pract 2013;19(5):402-11.
313. Ogundipe F, Kodadhala V, Ogundipe T, Mehari A, Gillum R. Disparities in Sepsis mortality by region, urbanization, and race in the USA: a multiple cause of death analysis. J Racial Ethn Health Disparities 2019;6(3):546-551.
314. Ogundipe F, Mehari A, Gillum R. Disparities in Sarcoidosis mortality by region, urbanization, and race in the United States: a multiple cause of death analysis. Am J Med 2019;132(9):1062-1068.e3.
315. Olie V, Fuhrman C, Chin F, et al. Time trends in pulmonary embolism mortality in France, 2000-2010. Thromb Res 2015;135(2):334-8.
316. Olson FE, Hammes LM, Shipley PW, Norris FD. A study of multiple causes of death in California. J Chronic Dis 1962;15(FEB):157-&.
317. Palumbo SA, Hennekens CH, Robishaw JD, Levine RS. Temporal trends and geographic variations in mortality rates from prescription Opioids: lessons from Florida and West Virginia. South Med J 2020;113(3):140-145.
318. Peden AE, Franklin RC, Mahony AJ, Scarr J, Barnsley PD. Using a retrospective cross-sectional study to analyse unintentional fatal drowning in Australia: ICD-10 coding-based methodologies verses actual deaths. BMJ Open 2017;7(12):e019407.
319. Perazzo H, Pacheco AG, De Boni R, et al. Age-standardized mortality rates related to cirrhosis in Brazil from 2000 to 2012: a nationwide analysis. Annals of Hepatology 2017;16(2):269-278.
320. Perkins BA, Flood JM, Danila R, et al. Unexplained deaths due to possibly infectious causes in the United States: defining the problem and designing surveillance and laboratory approaches. Emerg Infect Dis 1996;2(1):47-53.
321. Pickard AS, Jung E, Bartle B, Weiss KB, Lee TA. Coded cause of death and timing of COPD diagnosis. Copd 2009;6(1):41-7.
322. Piffaretti C, Moreno-Betancur M, Lamarche-Vadel A, Rey G. Quantifying cause-related mortality by weighting multiple causes of death. Bull World Health Organ 2016;94(12):870-879.
323. Polednak AP. US regional differences in death rates from depression. Soc Psychiatry Psychiatr Epidemiol 2012;47(12):1977-83.
324. Polednak AP. Recent decline in the U.S. death rate from myeloproliferative neoplasms, 1999-2006. Cancer Epidemiol 2012;36(2):133-6.
325. Prati C, Puyraveau M, Guillot X, Verhoeven F, Wendling D. Deaths associated with ankylosing spondylitis in France from 1969 to 2009. Journal of Rheumatology 2017;44(5):594-598.
326. Rahimi K, Duncan M, Pitcher A, Emdin CA, Goldacre MJ. Mortality from heart failure, acute myocardial infarction and other ischaemic heart disease in England and Oxford: a trend study of multiple-cause-coded death certification. Journal of Epidemiology & Community Health. 69(10):1000-5, 2015 Oct. 2015.
327. Rao C, Adair T, Bain C, Doi SA. Mortality from diabetic renal disease: a hidden epidemic. Eur J Public Health 2011;22(2):280-4.
328. Richardson DB. Use of multiple cause of death data in cancer mortality analyses. Am J Ind Med 2006;49(8):683-9.
329. Rinsky JL, Hoppin JA, Blair A, et al. Agricultural exposures and stroke mortality in the Agricultural Health Study. J Toxicol Environ Health A 2013;76(13):798-814.
330. Rio EM, Gallo PR, Reis AO. [Asthma mortality in the Municipality of São Paulo (1993-1995): analysis by multiple cause of death]. Cad Saude Publica 2003;19(5):1541-4.
331. Romon I, Jougla E, Balkau B, Fagot-Campagna A. The burden of diabetes-related mortality in France in 2002: an analysis using both underlying and multiple causes of death. Eur J Epidemiol 2008;23(5):327-34.
332. Rosenwax L, McNamara B, Zilkens R. A population-based retrospective cohort study comparing care for Western Australians with and without Alzheimer's disease in the last year of life. Health and Social Care in the Community 2009;17(1):36-44.
333. Sacks JJ, Helmick CG, Langmaid G. Deaths from arthritis and other rheumatic conditions, United States, 1979-1998. J Rheumatol 2004;31(9):1823-8.
334. Salonen J, Purokivi M, Bloigu R, Kaarteenaho R. Prognosis and causes of death of patients with acute exacerbation of fibrosing interstitial lung diseases. BMJ Open Respir Res 2020;7(1).
335. Santo AH. [Chickenpox-related mortality trends in the state of Sao Paulo, Brazil, 1985-2004: a multiple cause approach]. Rev Panam Salud Publica 2007;22(2):132-40.
336. Santo AH. Cysticercosis-related mortality in the State of Sao Paulo, Brazil, 1985-2004: A study using multiple causes of death. Cadernos De Saude Publica 2007;23(12):2917-2927.
337. Santo AH. [Epidemiological potential of multiple-cause-of-death data listed on death certificates, Brazil, 2003]. Rev Panam Salud Publica 2007;22(3):178-86.
338. Santo AH, Puech-Leao P, Krutman M. Trends in aortic aneurysm- and dissection-related mortality in the state of Sao Paulo, Brazil, 1985-2009: multiple-cause-of-death analysis. BMC Public Health 2012;12:859.
339. Santo AH. Sickle cell disease related mortality in Brazil, 2000-2018. Hematol Transfus Cell Ther 2020;44(2):177-185.
340. Sejvar JJ, Holman RC, Bresee JS, Kochanek KD, Schonberger LB. Amyotrophic lateral sclerosis mortality in the United States, 1979-2001. Neuroepidemiology 2005;25(3):144-52.
341. Selik RM, Rabkin CS. Cancer death rates associated with human immunodeficiency virus infection in the United States. J Natl Cancer Inst 1998;90(17):1300-1302.
342. Sheth SG, Krauss G, Krumholz A, Li G. Mortality in epilepsy: Driving fatalities vs other causes of death in patients with epilepsy. Neurology 2004;63(6):1002-1007.
343. Simmons R, Ireland G, Ijaz S, Ramsay M, Mandal S. Causes of death among persons diagnosed with hepatitis C infection in the pre- and post-DAA era in England: A record linkage study. J Viral Hepat 2019;26(7):873-880.
344. Sjogren H, Eriksson A, Ahlm K. Role of alcohol in unnatural deaths: a study of all deaths in Sweden. Alcoholism: Clinical & Experimental Research. 24(7):1050-6, 2000 Jul. 2000.
345. Sjogren H, Valverius P, Eriksson A. Gender differences in role of alcohol in fatal injury events. Eur J Public Health 2006;16(3):266-270.
346. Smestad C, Sandvik L, Celius EG. Excess mortality and cause of death in a cohort of Norwegian multiple sclerosis patients. Mult Scler 2009;15(11):1263-70.
347. Smith CA, Barnett E. Diabetes-related mortality among Mexican Americans, Puerto Ricans, and Cuban Americans in the United States. Rev Panam Salud Publica 2005;18(6):381-7.
348. Smithers-Sheedy H, Raynes-Greenow C, Badawi N, et al. Cytomegalovirus-related childhood mortality in Australia 1999-2011. J Paediatr Child Health 2015;51(9):901-5.
349. Snowdon DA, Phillips RL. Does a vegetarian diet reduce the occurrence of diabetes? Am J Public Health 1985;75(5):507-12.
350. Sondermeyer GL, Lee LA, Gilliss D, Vugia DJ. Coccidioidomycosis-associated deaths in California, 2000-2013. Public Health Rep 2016;131(4):531-5.
351. Steenland K, Steenland K, Nowlin S, et al. Use of multiple-cause mortality data in epidemiologic analyses: US rate and proportion files developed by the National Institute for Occupational Safety and Health and the National Cancer Institute. American Journal of Epidemiology 1992;136(7):855-862.
352. Steenland K, Attfield M, Mannejte A. Pooled analyses of renal disease mortality and crystalline silica exposure in three cohorts. Annals of Occupational Hygiene 2002;46:4-9.
353. Tardón AG, Zaplana Piñeiro J, Hernández Mejías R, Cueto Espinar A. Estudio de las Causas Múltiples de Defunción en Asturias, 1988. Gaceta Sanitaria 1993;7(35):78-85.
354. Thomas SL, Griffiths C, Smeeth L, Rooney C, Hall AJ. Burden of mortality associated with autoimmune diseases among females in the United Kingdom. Am J Public Health 2010;100(11):2279-87.
355. To T, Simatovic J, Zhu J, et al. Asthma deaths in a large provincial health system. A 10-year population-based study. Ann Am Thorac Soc 2014;11(8):1210-7.
356. Todd S, Barr S, Passmore AP. Cause of death in Alzheimer's disease: a cohort study. QJM 2013;106(8):747-53.
357. Trias-Llimos S, Martikainen P, Makela P, Janssen F. Comparison of different approaches for estimating age-specific alcohol-attributable mortality: The cases of France and Finland. PLoS One 2018;13(3):e0194478.
358. Tu EJ. Multiple cause-of-death analysis of hypertension-related mortality in New York State. Public Health Rep 1987;102(3):329-35.
359. Veazie M, Ayala C, Schieb L, et al. Trends and disparities in heart disease mortality among American Indians/Alaska Natives, 1990-2009. Am J Public Health 2014;104(SUPPL. 3):S359-S367.
360. Vierboom YC. Trends in alcohol-related mortality by educational attainment in the U.S., 2000-2017. Popul Res Policy Rev 2020;39(1):77-97.
361. Wall MM, Huang J, Oswald J, McCullen D. Factors associated with reporting multiple causes of death. BMC Med Res Methodol 2005;5(1):4.
362. Ward M, May P, Briggs R, et al. Linking death registration and survey data: Procedures and cohort profile for The Irish Longitudinal Study on Ageing. HRB Open Res 2020;3:43.
363. Westerling R. Small-area variation in multiple causes of death in Sweden - a comparison with underlying causes of death. International Journal of Epidemiology 1995;24(3):552-558.
364. Wier. Diabetes Mortality In Rhode Island: Comparing underlying cause of death versus any listed cause of death. 2008.
365. Wild SH, Bryden JR, Lee RJ, et al. Cancer, cardiovascular disease and diabetes mortality among women with a history of endometrial cancer. Br J Cancer 2007;96(11):1747-9.
366. Williams PT. Fifty-three year follow-up of coronary heart disease versus HDL2 and other lipoproteins in Gofman's Livermore Cohort. J Lipid Res 2012;53(2):266-72.
367. Wing S, Manton KG. A multiple cause of death analysis of hypertension-related mortality in North Carolina, 1968-1977. Am J Public Health 1981;71(8):823-30.
368. Wing S, Manton KG. The contribution of hypertension to mortality in the US: 1968, 1977. Am J Public Health. 73(2):140-4, 1983 Feb. 1983.
369. Wise ME, Sorvillo F. Hepatitis A--related mortality in California, 1989-2000: analysis of multiple cause-coded death data. Am J Public Health 2005;95(5):900-5.
370. Wise M, Bialek S, Finelli L, Bell BP, Sorvillo F. Changing trends in hepatitis C-related mortality in the United States, 1995-2004. Hepatology 2008;47(4):1128-35.
371. Xie SH, Chen H, Lagergren J. Causes of death in patients diagnosed with gastric adenocarcinoma in Sweden, 1970-2014: A population-based study. Cancer Sci 2020;111(7):2451-2459.
372. Yamamura M, Santos-Neto M, dos Santos RAN, et al. Epidemiological characteristics of cases of death from tuberculosis and vulnerable territories. Revista Latino-Americana de Enfermagem 2015;23(5):910-918.
373. Yang Q, Khoury MJ, Mannino D. Trends and patterns of mortality associated with birth defects and genetic diseases in the United States, 1979-1992: an analysis of multiple-cause mortality data. Genet Epidemiol 1997;14(5):493-505.
374. Yoon YH, Yi HY, Thomson PC. Alcohol-related and viral hepatitis C-related cirrhosis mortality among Hispanic subgroups in the United States, 2000-2004. Alcohol Clin Exp Res 2011;35(2):240-9.
375. Zhu M, Li J, Li Z, et al. Mortality rates and the causes of death related to diabetes mellitus in Shanghai Songjiang District: an 11-year retrospective analysis of death certificates. BMC Endocr Disord 2015;15:45.
376. Zilkens RR, Spilsbury K, Bruce DG, Semmens JB. Linkage of hospital and death records increased identification of dementia cases and death rate estimates. Neuroepidemiology 2009;32(1):61-9.
377. Zoppini G, Fedeli U, Schievano E, et al. Mortality from infectious diseases in diabetes. Nutr Metab Cardiovasc Dis 2018;28(5):444-450.
378. Adair T, Rao C. Changes in certification of diabetes with cardiovascular diseases increased reported diabetes mortality in Australia and the United States. J Clin Epidemiol 2010;63(2):199-204.
379. Axtell CD, Ward EM, McCabe GP, et al. Underlying and multiple cause mortality in a cohort of workers exposed to aromatic amines. Am J Ind Med 1998;34(5):506-11.
380. Ball LB, Macdonald SC, Mott JA, Etzel RA. Carbon monoxide-related injury estimation using ICD-coded data: methodologic implications for public health surveillance. Arch Environ Occup Health 2005;60(3):119-27.
381. Batty GD, Gale CR, Kivimaki M, Bell S. Assessment of Relative Utility of Underlying vs Contributory Causes of Death. JAMA Netw Open 2019;2(7):e198024.
382. Bohm MKC, H. B. Nonmedical use of prescription Opioids, heroin use, injection drug use, and overdose mortality in U.S. adolescents. J Stud Alcohol Drugs 2020.
383. Cano M, Oh S, Salas-Wright CP, Vaughn MG. Cocaine use and overdose mortality in the United States: Evidence from two national data sources, 2002-2018. Drug Alcohol Depend 2020;214:108148.
384. Carrillo-Larco RM, Bernabe-Ortiz A. A divergence between underlying and final causes of death in selected conditions: an analysis of death registries in Peru. PeerJ 2018;6:e5948.
385. Chamblee RF, Evans MC, Patten DG, Pearce JS. Injuries causing death: Their nature, external causes, and associated diseases. J Safety Res 1983;14(1):21-35.
386. Chang CY, Lu TH, Cheng TJ. Trends in reporting injury as a cause of death among people with epilepsy in the U.S., 1981-2010. Seizure 2014;23(10):836-43.
387. Chitty KM, Schumann JL, Moran LL, et al. Reporting of alcohol as a contributor to death in Australian national suicide statistics and its relationship to post-mortem alcohol concentrations. Addiction.
388. Fazito E, Vasconcelos AM, Pereira MG, Rezende DF. Trends in non-AIDS-related causes of death among adults with HIV/AIDS, Brazil, 1999 to 2010. Cad Saude Publica 2013;29(8):1644-53.
389. Fedeli U, Avossa F, Guzzinati S, Bovo E, Saugo M. Trends in mortality from chronic liver disease. Ann Epidemiol 2014;24(7):522-6.
390. Fingerhut LA, Warner M. The ICD-10 injury mortality diagnosis matrix. Inj Prev 2006;12(1):24-9.
391. Fuller JH, Elford J, Goldblatt P, Adelstein AM. Diabetes mortality: new light on an underestimated public health problem. Diabetologia 1983;24(5):336-41.
392. Hansen J, Asberg S, Kumlien E, Zelano J. Cause of death in patients with poststroke epilepsy: Results from a nationwide cohort study. PLoS One 2017;12(4):e0174659.
393. Jerschow E, Lin RY, Scaperotti MM, McGinn AP. Fatal anaphylaxis in the United States, 1999-2010: temporal patterns and demographic associations. J Allergy Clin Immunol 2014;134(6):1318-1328 e7.
394. Kramarow E, Warner M, Chen LH. Food-related choking deaths among the elderly. Injury Prevention 2014;20(3):200-203.
395. Krapfl HR, Gohdes DM, Croft JB. Racial and ethnic differences in premature heart disease deaths in New Mexico: what is the role of diabetes? Ethn Dis 2006;16(1):85-8.
396. Kravitz-Wirtz N, Davis CS, Ponicki WR, et al. Association of Medicaid expansion With Opioid overdose mortality in the United States. JAMA Netw Open 2020;3(1):e1919066.
397. Lahti RA, Korpi H, Vuori E. Blood-positive illicit-drug findings: implications for cause-of-death certification, classification and coding. Forensic Sci Int 2009;187(1-3):14-8.
398. Lanska DJ, Lanska M, Lavine L, Schoenberg BS. Conditions associated with Huntington's Disease at death: A case-control study. Archives of Neurology 1988;45(8):878-880.
399. Mackenbach JP, Kunst AE, Lautenbach H, Oei YB, Bijlsma F. Gains in life expectancy after elimination of major causes of death: Revised estimates taking into account the effect of competing causes. J Epidemiol Community Health 1999;53(1):32-37.
400. Manton KG, Tolley DH, Poss SS. Life table techniques for multiple-cause mortality. Demography 1976;13(4):541-64.
401. Manton KG, Sharon SP. Effects of dependency among causes of death from elimination life table strategies. Demography 1979;16(2):313-327.
402. Manton KG, Stallard E, Poss SS. Estimates of U.S. multiple cause life tables. Demography 1980;17(1):85-102.
403. Martikainen P, Makela P, Peltonen R, Myrskyla M. Income differences in life expectancy: the changing contribution of harmful consumption of alcohol and smoking. Epidemiology. 25(2):182-90, 2014 Mar. 2014.
404. McDonnell WF, Nishino-Ishikawa N, Petersen FF, Chen LH, Abbey DE. Relationships of mortality with the fine and coarse fractions of long-term ambient PM10 concentrations in nonsmokers. Journal of Exposure Analysis and Environmental Epidemiology. 10(5):427-36, 2000 Sep-Oct. 2000.
405. Miller TR, Swedler DI, Lawrence BA, et al. Incidence and lethality of suicidal overdoses by drug class. JAMA Netw Open 2020;3(3):e200607.
406. Moussa MA. Analysis of underlying and multiple-cause mortality data: the life table methods. Comput Methods Programs Biomed 1987;24(1):3-19.
407. Pacheco AG, Saraceni V, Tuboi SH, et al. Estimating the extent of underreporting of mortality among HIV-Infected individuals in Rio de Janeiro, Brazil. AIDS Research and Human Retroviruses 2011;27(1):25-28.
408. Pinault L, Brauer M, Crouse DL, et al. Diabetes status and susceptibility to the effects of PM2.5 exposure on cardiovascular mortality in a national Canadian cohort. Epidemiology 2018;29(6).
409. Quast TC. Potential undercounting of overdose deaths caused by specific drugs in vital statistics data: An analysis of Florida. Drug Alcohol Depend 2020;207:107807.
410. Quinones A, Lobach I, Maduro GA, Jr., Smilowitz NR, Reynolds HR. Diabetes and ischemic heart disease death in people age 25-54: a multiple-cause-of-death analysis based on over 400 000 deaths from 1990 to 2008 in New York City. Clin Cardiol 2015;38(2):114-20.
411. Ramos AN, Jr., Matida LH, Hearst N, Heukelbach J. Mortality in Brazilian children with HIV/AIDS: the role of non-AIDS-related conditions after highly active antiretroviral therapy introduction. AIDS Patient Care STDS 2011;25(12):713-8.
412. Redelings MDL, N. E.; Sorvillo, F. Pressure ulcers: more lethal than we thought? Adv Skin Wound Care 2005.
413. Redelings MD, Sorvillo F, Simon P. A comparison of underlying cause and multiple causes of death: US vital statistics, 2000-2001. Epidemiology 2006;17(1):100-3.
414. Redelings MD, Wise M, Sorvillo F. Using multiple cause-of-death data to investigate associations and causality between conditions listed on the death certificate. Am J Epidemiol 2007;166(1):104-8.
415. Rezende EM, Sampaio IB, Ishitani LH. [Multiple causes of death due to non-communicable diseases: a multidimensional analysis]. Cad Saude Publica 2004;20(5):1223-31.
416. Rezende EM, Sampaio BM, Ishitani LH, Martins EF, Vilella Lde C. [Mortality of malnourished elderly in Belo Horizonte, Minas Gerais State, Brazil: a multidimensional analysis focusing on multiple causes of death]. Cad Saude Publica 2010;26(6):1109-21.
417. Riihimaki M, Thomsen H, Brandt A, Sundquist J, Hemminki K. What do prostate cancer patients die of? Oncologist. 16(2):175-81, 2011. 2011.
418. Rockett IR, Wang S, Lian Y, Stack S. Suicide-associated comorbidity among US males and females: a multiple cause-of-death analysis. Inj Prev 2007;13(5):311-5.
419. Rockett IR, Lian Y, Stack S, Ducatman AM, Wang S. Discrepant comorbidity between minority and white suicides: a national multiple cause-of-death analysis. BMC Psychiatry 2009;9:10.
420. Salazar A, Moreno S, De Sola H, et al. The evolution of opioid-related mortality and potential years of life lost in Spain from 2008 to 2017: differences between Spain and the United States. Curr Med Res Opin 2020;36(2):285-291.
421. Selik RM, Chu SY, Ward JW. Trends in infectious diseases and cancers among persons dying of HIV infection in the United States from 1987 to 1992. Ann Intern Med 1995;123(12):933-6.
422. Selik RM, Byers Jr RH, Dworkin MS. Trends in diseases reported on U.S. death certificates that mentioned HIV infection, 1987-1999. Journal of Acquired Immune Deficiency Syndromes 2002;29(4):378-387.
423. Seuc AH, Fernandez-Gonzalez L, Mirabal M. Comparative Disease Assessment: a multi-causal approach for estimating the burden of mortality. Journal of Public Health 2020;30(3):665-673.
424. Shah NA, Abate MA, Smith MJ, et al. Characteristics of alprazolam-related deaths compiled by a centralized state medical examiner. Am J Addict 2012;21 Suppl 1:S27-34.
425. Sosin DM, Sniezek JE, Waxweiler RJ. Trends in death associated with traumatic brain injury, 1979 through 1992. Success and failure. Jama 1995;273(22):1778-80.
426. Thomas BM, Starr JM, Whalley LJ. Death certification in treated cases of presenile Alzheimer's disease and vascular dementia in Scotland. Age Ageing 1997;26(5):401-406.
427. Tolley HD, Manton KG, Poss SS. A linear models application of competing risks to multiple causes of death. Biometrics 1978;34(4):581-91.
428. Warner M, Paulozzi LJ, Nolte KB, Davis GG, Nelson LS. State variation in certifying manner of death and drugs involved in drug intoxication deaths. Academic Forensic Pathology 2013;3(2):231-237.
429. Weiner LB, MT; McAvoy, GH et al. Use of multiple causes in the classification of deaths from cardiovascular-renal disease. Am J Public Health. 14(2):100-5, 1924 Feb. 1955.
430. Whelton PK, Goldblatt P. An investigation of the relationship between stomach cancer and cerebrovascular disease: evidence for and against the salt hypothesis. Am J Epidemiol 1982;115(3):418-27.
431. You HS, Ha J, Kang CY, et al. Regional variation in states' naloxone accessibility laws in association with opioid overdose death rates-Observational study (STROBE compliant). Medicine (Baltimore) 2020;99(22):e20033.
432. Breger TL, Edwards JK, Cole SR, et al. Estimating a set of mortality risk functions with multiple contributing causes of death. Epidemiology 2020;31(5):704-712.
433. Hassanzadeh HR, Sha Y, Wang MD. DeepDeath: Learning to predict the underlying cause of death with Big Data. 2017 39th Annual International Conference of the IEEE Engineering in Medicine and Biology Society 2017:3373-3376.
434. Jiang H, Wu H, Wang MD. Causes of death in the United States, 1999 to 2014. 2017 IEEE EMBS International Conference on Biomedical and Health Informatics, BHI 2017 2017:177-180.
